# Possible future waves of SARS-CoV-2 infection generated by variants of concern with a range of characteristics

**DOI:** 10.1101/2021.06.07.21258476

**Authors:** Louise Dyson, Edward M. Hill, Sam Moore, Jacob Curran-Sebastian, Michael J. Tildesley, Katrina A Lythgoe, Thomas House, Lorenzo Pellis, Matt J. Keeling

## Abstract

Viral reproduction of SARS-CoV-2 provides opportunities for the acquisition of advantageous mutations, altering viral transmissibility, disease severity, and/or allowing escape from natural or vaccine-derived immunity. We use three mathematical models: a parsimonious deterministic model with homogeneous mixing; an age-structured model; and a stochastic importation model to investigate the effect of potential variants of concern (VOCs). Calibrating to the situation in England in May 2021, we find epidemiological trajectories for putative VOCs are wide-ranging and dependent on their transmissibility, immune escape capability, and the introduction timing of a postulated VOC-targeted vaccine. We demonstrate that a VOC with a substantial transmission advantage over resident variants, or with immune escape properties, can generate a wave of infections and hospitalisations comparable to the winter 2020-2021 wave. Moreover, a variant that is less transmissible, but shows partial immune-escape could provoke a wave of infection that would not be revealed until control measures are further relaxed.

## Introduction

Since the SARS-CoV-2 virus was first identified in humans in late 2019, the resulting global pandemic has caused, as of 23rd July 2021, over 190 million confirmed COVID-19 cases and above 4.1 million reported deaths with COVID-19 disease [1]. As the pandemic continues globally, high SARS-CoV-2 incidence rates act to increase the risk of the virus acquiring additional advantageous mutations [2], potentially altering transmissibility, severity and escape from natural or vaccine-derived immunity.

The number of countries reporting variants causing concern continues to increase [1]. One such SARS-CoV-2 variant is PANGO lineage B.1.1.7 [3], with WHO label ‘Alpha’ [4]. This variant was first detected in southeast England, with the earliest sequenced B.1.1.7 samples collected in September 2020 [5]. The B.1.1.7 variant was designated a variant of concern (VOC) in the UK on 18th December 2020 [6]. There is a consensus across multiple statistical and mechanistic modelling approaches that the B.1.1.7 variant has a substantial transmission advantage over preexisting variants, with estimates ranging between 40 and 80% more transmissible than previously-circulating variants [7–11]. Furthermore, matched cohort studies also suggest the B.1.1.7 variant is associated with higher mortality compared to preexisting variants at a population level [12–14], although there appears to be no significant difference in mortality for cases we already know to be severe enough as to require hospitalisation [15].

Subsequently, the B.1.617.2 PANGO lineage (with WHO label ‘Delta’ [4]), a variant initially prevalent in India [16], was designated a VOC in the UK on 6th May 2021 due to it being assessed to have “at least equivalent transmissibility to B.1.1.7 based on available data (with moderate confidence)” [17]. A week later, on 13th May 2021, this assessment was revised to “high confidence” [18]. The continued growth of B.1.617.2 relative to B.1.1.7 observed in the UK is indicative of a substantial transmission advantage [19].

Novel variants of COVID-19 that substantially evade vaccine or naturally-acquired immunity may pose a much bigger threat than those that somewhat increase overall transmissibility, reducing the efficacy of vaccines and enabling higher rates of re-infection. There is apprehension that as countries with high vaccine coverage begin to relax nonpharmaceutical interventions (NPIs), variants may be revealed within-country, or be imported, that escape existing immunity, thereby causing new waves of infection. Notably, initial evidence for the B.1.351 variant (with WHO label ‘Beta’ [4]) suggests potential immune escape; B.1.351 was first detected in South Africa in October 2020 [20] and was designated a VOC in the UK on 24th December 2020. Collective findings from neutralisation experiments, vaccine clinical trials and observational studies of population-level surveillance data indicate that B.1.351 can evade natural immunity from previous infection [21], and the two prominently used SARS-CoV-2 vaccines in the UK, the AstraZeneca (AZ) and Pfizer-BioNTech (Pfizer) vaccines, likely have reduced efficacy against B.1.351 [22–25]. It has been suggested that immune evasion explains the growth of B.1.351 in some regions of France [26]. There has also been concern that the variant P.1 (with WHO label ‘Gamma’ [4]), first reported in Manaus, Brazil, in December 2020, can evade immunity; a large secondary wave of infection occurred in Manaus despite high-levels of pre-existing immunity due to a previous large wave of infection [27], although, neutralisation experiments have been more equivocal [28]. Perhaps most worrying is the potential for the emergence of variants that are both highly transmissible and harbour immune-escape mutations. Hence, B.1.1.7 lineages that also have the E484K mutation, which is associated with reduced neutralisation from antibodies, were designated a VOC in its own right in the UK on 5th February 2021 [29].

A range of different VOCs are found in genomically sequenced specimens in England. Of those reported by 31st May 2021 (noting that delays between specimen collection and sequencing can extend to up to three weeks), there had been 846 genomically sequenced samples of B.1.351, 151 of P.1, 43 of B.1.1.7 with E484K and 9,426 of B.1.617.2 variant cases (excluding variant cases not linked to a known COVID-19 case or with provisional sequencing/genotyping results) [30]. We remark that the frequency of variants among sequenced samples may not be representative of variant frequencies more broadly due to non-random selection of samples sent for sequencing.

The infectious disease dynamics of SARS-CoV-2 result from a complex interaction between the circulation/presence of multiple variants, vaccination, NPI policy and adherence. Mathematical modelling approaches are an avenue for testing sensitivity of these dynamics to underlying assumptions and conveying uncertainty, with the caveat that models must balance biological realism with mathematical and computational tractability and parameter identifiability [31]. Models have previously demonstrated their usefulness as a tool offering insights on the dynamics of pathogens with multiple lineages [32, 33]. At the original time of writing (May 2021) there was burgeoning interest in modelling to explore the effects of VOCs on the trajectory of the SARS-CoV-2 epidemic. One such paper used a deterministic compartmental model to simulate the impact of the potential introduction of the more transmissible variant, B.1.1.7, into a Colombian population in which previous strains were dominant [34]. The authors considered the effect on prevalence of hospitalisation and deaths, and concluded that the introduction of such a variant would necessitate increased NPIs and an increased pace of vaccinations, though the potential immune escape characteristics of a VOC were not explored. Another example study, considering the spread of the B.1.1.7 variant in Ontario, Canada, devised a two-strain mathematical framework to model both a resident and a mutant-type viral population to estimate the time at which a mutant variant is able to take over a resident-type strain during an emerging infectious disease outbreak [35].

In this study, we use three mathematical models of novel SARS-CoV-2 variant dynamics to evaluate the drivers, and the likely timescales, of SARS-CoV-2 VOC epidemics in England. We demonstrate that a VOC can cause subsequent epidemic outbreaks comparable in magnitude to earlier waves in the pandemic if it possesses either a large transmission advantage over the existing resident variants, or the ability to evade immunity (either infection- or vaccine-derived). Further, even when a novel variant is less transmissible than the locally resident variants, immune escape can lead to a marked wave of infection and consequential hospitalisations. In addition, the reduced transmissibility of such a VOC can allow it to remain difficult to detect until NPIs are reduced. Finally, we explore the relative timing of VOC-targeted vaccines versus the establishment of community transmission of an emergent VOC, and show a multitude of projected possibilities, demonstrating the need to remain attentive to all potential scenarios.

## Results

We investigate two ways in which variants may be concerning: either that they may be more transmissible than the resident variants; or that they may evade immunity (infection- or vaccine-derived). While there are indications of immune escape for some particular variants [23, 24], the extent to which these variants evade immunity in vivo is uncertain [27]. We therefore begin by exploring parameter space using a parsimonious deterministic model with simple homogeneous mixing. While such a parsimonious model is useful for exploring parameter space and understanding the essential dynamics, it is unsuited to understanding how infection may be translated into disease burden. We therefore extend our analysis using a more complex age-structured model: firstly to ensure that the simplifications made for the parsimonious model do not have a large effect on the dynamics and secondly to investigate how the infections seen in the parsimonious model may translate into hospitalisations. In particular, as the roll-out of vaccinations progresses, we expect the proportion of infections that result in hospitalisations or death to decrease, reducing the burden of large numbers of infections. Finally, using a Gillespie stochastic simulation we explore how the timing of the introduction of a putative VOC-targeted vaccine and the rate of VOC introductions into the population impact the trajectory of the epidemic. See the Methods for further details on the three models used to perform our analyses.

We considered six representative potential VOCs, specifying relative transmissibility versus resident variants (with resident variants in our context referring to the period in England when the Alpha/B.1.1.7 variant was predominant) and immune escape properties (Table 1): VOC MT – More transmissible, no immune escape; VOC E – equally transmissible, (partial) immune escape to vaccination and prior infection; VOC LT+E – Less Transmissible, (partial) immune escape to vaccination and prior infection; VOC Ev – Equally transmissible, (partial) immune escape to vaccination only; VOC Ei – Equally transmissible, (partial) immune escape to prior infection only; VOC E+LH – as VOC E, but with no immune escape to hospitalisation. Results for VOCs Ev and Ei are presented in the Supplementary Information. While these scenarios focused on VOCs that had either an advantage in terms of transmissibility or to escape previously acquired immunity (but not both), our sensitivity analyses considered VOCs possessing a combination of both advantages.

**Table 1:** Transmissibility and infection immune escape properties for putative VOCs. In the main analysis we consider four VOCs (VOC MT, VOC E, VOC LT+E, VOC E+LH), with results for two additional VOCs (VOC Ev and VOC Ei) presented in the Supplementary Information. For those previously infected by either the resident variant or the VOC, we assumed the prior-infection efficacies towards the other variant were identical. Note that in the age-structured SARS-CoV-2 transmission model we also applied efficacy scalings upon both symptomatic disease and hospitalisations (severe disease).

| Scenario | Description | Relative transmissibility | Proportional vaccine efficacy | Proportional prior-infection efficacy |
| --- | --- | --- | --- | --- |
| VOC MT | More transmissible, no immune escape | 1.5 | 1 | 1 |
| VOC E | Equally transmissible, immune escape to vaccination and prior infection | 1 | 0.75 | 0.75 |
| VOC E+LH | As VOC E, except efficacies against hospitalisations unadjusted | 1 | 0.75* | 0.75* |
| VOC LT+E | Less transmissible, immune escape to vaccination and prior infection | 0.8 | 0.75 | 0.75 |
| VOC Ev | Equally transmissible, immune escape to vaccination only | 1 | 0.75 | 1 |
| VOC Ei | Equally transmissible, immune escape to prior infection only | 1 | 1 | 0.75 |

\* VOC E+LH displays full efficacy against hospitalisations.

### Effects of potential variants on resultant waves of SARS-CoV-2 infection

In the absence of any introductions of other variants and assuming the continuation of the relaxation roadmap to Step 4 from 21st June 2021, the parsimonious SARS-CoV-2 transmission model gave a small wave of infection for currently-circulating variants (primarily B.1.1.7) spanning the second half of 2021 with a peak infectious prevalence of approximately 0.5% (Figure 1a, black line). This is in broad agreement with contemporaneous modelling of the roadmap relaxations [38].

**Figure 1:**
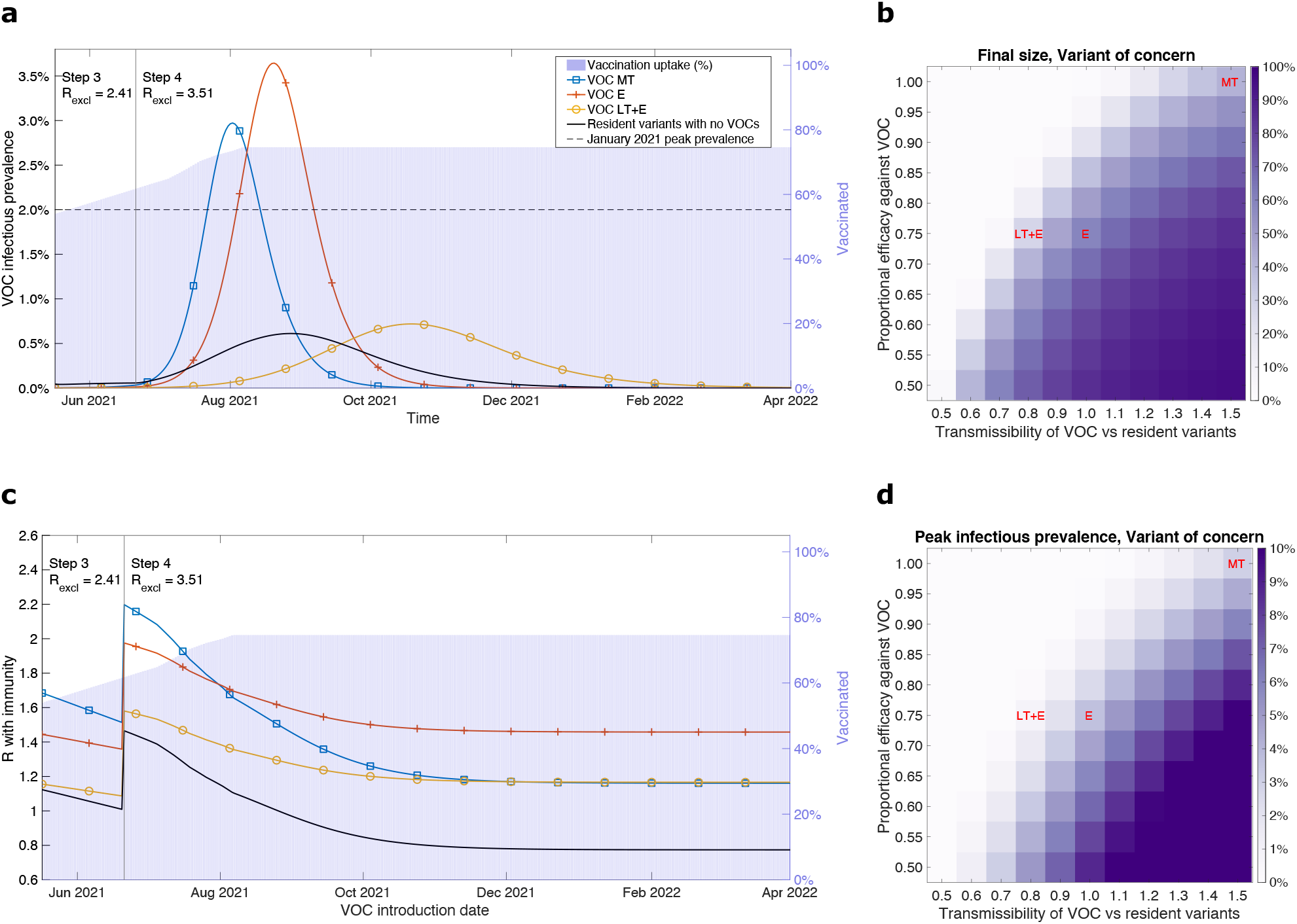
Infection burden for illustrative VOC scenarios, produced using the parsimonious SARS-CoV-2 transmission model. We considered three putative VOCs with differing transmissibility and immune escape characteristics: more transmissible (VOC MT, blue line with square markers), equal transmissibility with immune escape (VOC E, orange line with plus sign markers), less transmissible with immune escape (VOC LT+E, yellow line with circle markers), and resident variants alone in the absence of any VOC being introduced (black line with no markers). Additionally, in panels **a&c** we represent the vaccine uptake in the population through time via background shading, the transition time into Step 4 of the relaxation roadmap by the vertical solid line and we state the assumed R excluding immunity values for resident variants (*R*_excl_) throughout Steps 3 and 4, respectively. **a** VOC infectious prevalence over time. In each scenario, alongside resident variants, we introduced one of the VOCs on 17th May 2021 with 2,000 initial infecteds. **c** Instantaneous R of a VOC accounting for population-level immunity (y axis) calculated at the time of its introduction (x axis). For the ‘Resident variants with no VOCs’ scenario the displayed profile corresponds to the instantaneous R with immunity of resident variants. In panels **b&d** we explore the sensitivity of three epidemiological outcomes to the relative transmissibility of the VOC compared to resident variant and the proportional efficacy (vaccine and natural-immunity) against the VOC: **b** outbreak final size; **d** peak in VOC infection cases.

On the other hand, VOCs can lead to waves of infection beyond what we would expect from the resident variants. The introduction of a variant that was 1.5 times more transmissible than resident variants (VOC MT) resulted in a surge of infection peaking in August 2021. Additionally, the peak exceeded the estimated peak prevalence during the January 2021 wave in England as estimated from the ONS infection survey [50] (Figure 1a, blue line with square markers).

Similarly VOC E, which, whilst no more transmissible than resident variants, had a degree of immune escape from vaccination-derived immunity or prior infection (25% reduction compared to resident variants), also provoked a considerable wave of VOC infections. Compared to the more transmissible VOC MT, the epidemic wave was lagged by a month, with a peak in infectious prevalence in excess of the estimated peak prevalence during the January 2021 wave (Figure 1a, orange line with plus sign markers).

VOCs that had only one component of immune escape, to either vaccination only or prior infection only, displayed shallower and broader epidemic waves compared to VOC E. We found that VOC Ei (immune escape to prior infection only) peaked a month late with a higher magnitude and had a longer epidemic tail than VOC Ev (immune escape to vaccination only, see Supplementary Fig. 5).

A variant that was less transmissible than the resident variants but had immune escape attributes, VOC LT+E, could give rise to an elongated epidemic that was flatter, and more delayed, than VOC E (Figure 1a). These dynamics were a consequence of the relative growth of the two variants depending on a combination of relative transmissibility and relative immunity.

The trajectory of the initial resident variant was similar soon after the introduction of any of the VOCs. Increases in VOC infections then translated into increased immunity against resident variant infections, which resulted in trajectories diverging by late July 2021. As a consequence, the VOCs with large resultant infection waves (VOC MT and VOC E) coincided with a shallower, earlier peak in resident variant infectious prevalence and a shortened outbreak duration for resident variants (Supplementary Fig. 6).

The relationship between the temporal profiles of VOC and resident variant infectious prevalence were also reflected in the speed of replacement of the resident variants by the VOC. Our more transmissible variant, VOC MT, encompassed 50% of cases less than two months post-introduction and approached 100% of cases within three months (August 2021), whereas in the VOC scenario with a less transmissible variant with immune escape, VOC LT+E, it took roughly four months after being introduced (during September 2021) for the VOC to constitute 50% of cases (Supplementary Fig. 7).

Both the outbreak size and peak in infectious prevalence for VOCs were sensitive to the transmissibility and ability to evade existing immunity (Figure 1b,d). We found a highly transmissible VOC, 1.5 times more transmissible than resident variants, that also had a great ability to evade prior-infection and vaccine-derived immunity (proportional immune efficacy against the VOC of 0.5), could cause outbreaks infecting the majority of the population and attain a peak infectious prevalence approaching 10%. On the other hand, outbreaks were generally not sustained for VOCs that had a combination of being less transmissible than resident variants with only minor evasion of infection- and vaccine-derived immunity.

As time goes on, both the immunity of the population (via vaccinations and infections with resident strains) and the level of NPIs change, leading to different dynamics depending on when a VOC is introduced. Different types of VOCs have more advantage at different dates of introduction, reflected in the value of R with immunity (also referred to as effective R, with notation 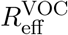) at the time of introduction. While more transmissible variants (such as VOC MT) would attain their highest instantaneous R with immunity estimate (at the time of introduction) if the date of introduction aligned with the date of moving to Step 4 of the relaxation roadmap on 21st June 2021, that advantage is degraded over time as the population builds immunity (Figure 1c, blue line with square markers). Variants that have a degree of immune escape then gain greater relative advantage if the VOC was introduced at a later time (Figure 1c, VOC E and VOC LT+E). Notably, introducing either VOC MT or VOC LT+E from November 2021 or later resulted in matched R with immunity values, plateauing at approximately 1.2. Conversely, before Step 4 of the relaxation roadmap occurs, VOC LT+E may be quite indistinguishable from resident variants.

These facets were borne out by comparing outbreak size and infectious case peak summary statistics; an introduction of either VOC MT and VOC LT+E in late 2021 gave similar epidemic trajectories. Further, VOC MT being introduced in late 2021, rather than 17th May 2021, resulted in a greater than three-fold reduction in outbreak size and peak infectious prevalence. For VOC E, quantitatively the impacts of a later introduction date were less marked. In particular, a later introduction date led to only a small decrease in the outbreak size from approximately 60% (for an introduction date of 17th May 2021) to 50% (for an introduction date in August 2021) of the population, respectively (Supplementary Fig. 8, panels a-c). Contrarily, there was less variability in the outbreak summary statistics for resident variants, irrespective of the VOC that was introduced into the transmission dynamics (Supplementary Fig. 8, panels d-f).

Furthermore, we sought to determine what characteristics a VOC needed to possess to both spread through the population (i.e. 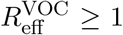) and outcompete resident variants (i.e 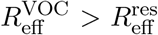). If the VOC was introduced on 17th May 2021, immune escape was not necessarily required if the VOC was more transmissible than resident variants (Supplementary Fig. 9a). For VOCs that were less transmissible than resident variants, a 10% decrement in relative transmissibility could be roughly offset by a 10% decrement in the proportional efficacy of immunity against the VOC. For later VOC introduction dates of 1st August 2021 (Supplementary Fig. 9b) and 1st November 2021 (Supplementary Fig. 9c), higher relative transmissibilities were required for VOCs that did not have much immune escape (proportional efficacy against the VOC of 0.9 and above), but lower relative transmissibilities could be successful for VOCs that had high immune escape (proportional efficacy against the VOC of 0.75 and below).

Though individuals may develop immunity due to prior infection or vaccination, it can be imperfect and breakthrough infections may occur. In these circumstances, the immune response could still cause a reduction in the onward transmission of the virus. Including a degree of transmission blocking (by either 25% or 50%) for those suffering breakthrough infection resulted in a reduction in any resultant wave of VOC infections and delayed the peak of the epidemic wave (Supplementary Fig. 10). For VOC E, in particular, a 50% transmission blocking effect shifted the epidemic wave into late 2021 and early 2022, while reducing the peak in infection to less than a third compared to no transmission blocking. Transmission blocking from vaccinations also reduced the maximum attained effective R over the course of the outbreak (Supplementary Fig. 11). For the less transmissible VOC LT+E, a 25% transmission blocking effect was sufficient to prevent any further substantial outbreak, with effective R kept below 1.5 throughout.

### VOC-caused hospitalisation burden depends sensitively on VOC attributes

Since the parsimonious model did not include age structure, it was unable to incorporate correlations between individuals that are prioritised for vaccination, those that contribute most to transmission and those most susceptible to severe disease, whose outcomes may require hospital treatment. In the UK older age groups were prioritised for vaccination, representing a population that are most at risk of severe disease, but contribute least to onward infection. To investigate the effect of this correlation, and include reductions in the severity of cases due to vaccinations, we turned to a more complex age-structured model. As before, we considered a range of potential effects on transmissibility and immunity, either from prior infections or from vaccinations. In addition, we included the potential effect of a (partial) immune-escape variant that assumed no reduction in vaccine-derived efficacy against hospitalisation (VOC E+LH).

Our results for the age-structured model broadly agreed with the parsimonious model in terms of qualitative patterns between the illustrative VOC scenarios. When the relaxation roadmap in England proceeded at the earliest stipulated dates, both a variant that was 50% more transmissible than the resident variant with no immune escape attributes (VOC MT) or an equally transmissible VOC with a reduction in efficacy from infection- and vaccine-derived immunity (VOC E) were sufficient to see a substantial outbreak. Furthermore, these cases can result in appreciable hospital admissions, which may exceed the daily peak attained during January 2021 of 3,700 admissions per day across England (Figure 2a).

**Figure 2:**
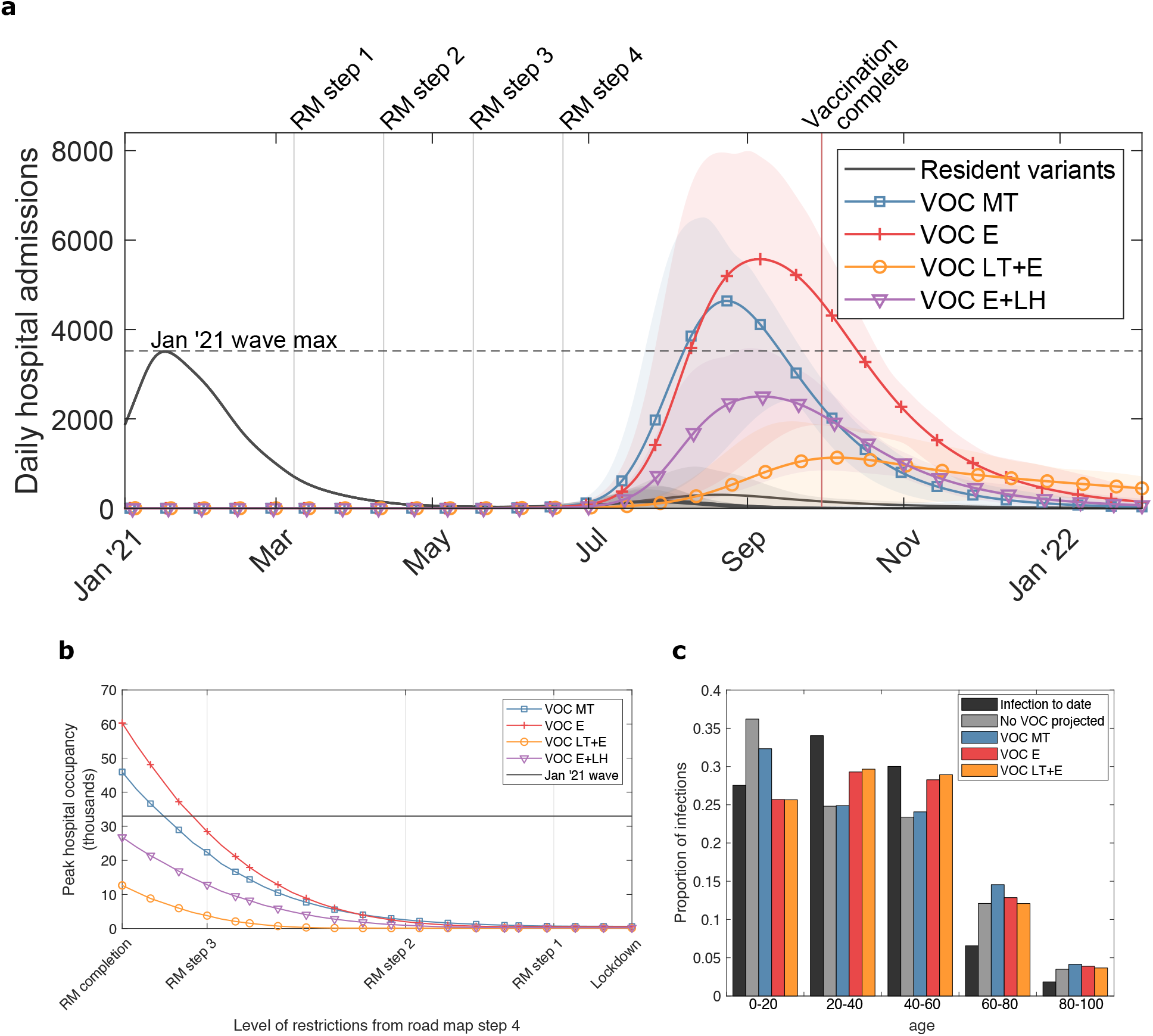
Estimated COVID-19 hospitalisations, using the age-structured SARS-CoV-2 transmission model, across the illustrative VOC scenarios. We considered four putative VOCs with differing transmissibility, severity and immune escape characteristics: more transmissible (VOC MT, blue, square markers), equal transmissibility with immune escape (VOC E, orange, plus sign markers), less transmissible with vaccine immune escape (VOC LT+E, yellow, circle markers) and equal transmissibility with the same immune escape properties of VOC E with the exception of of a lesser reduction in vaccine-derived efficacy against hospitalisation (VOC E+LH, purple, inverted triangle markers). **a** Time series of daily hospital admissions (thousands). Solid lines show the mean at each timepoint and the shaded ribbons the 95% prediction intervals. The dashed horizontal line denotes the peak in daily hospital admissions in England during the January 2021 wave. Vertical grey lines give the timing of each Step of the relaxation roadmap, with Step 4 being placed at the earliest stipulated date that it may begin (21st June 2021). The vertical light red line corresponds to the projected date under our vaccine roll-out speed assumption where all those in the adult population in England who accept the vaccine would have received two doses. **b** Relationship between mean peak hospital occupancy with VOC (thousands) and the level of NPIs towards the population following Step 4 of the relaxation roadmap. **c** Age distribution of infections from the historical data (black bars) alongside the projected distributions for the resident variant in the absence of any VOCs (grey bars) and each VOC scenario (VOC MT: blue bars; VOC E: red bars; VOC LT+E: orange bars).

It is hoped that, even when the effect of current vaccines and natural immunity in preventing infection are significantly compromised, they may still be effective in preventing severe symptom effects. Nevertheless, when both vaccination and previous infection are equally effective at preventing hospitalisations from both VOC and resident variants (VOC E+LH), we retain a large wave of resultant hospitalisations generated by the variant, though the central trajectory is brought below the peak level of daily hospital admissions during the January 2021 wave (Figure 2a, purple line).

The burden of cases with severe disease being admitted to hospital could be diminished with prolonged use of NPIs. The stringency of these NPIs would depend on the characteristics of the variant, though the non-COVID harms would also need consideration. Irrespective of the level of restrictions retained in Step 4 of the roadmap, a high vaccine efficacy against severe disease reduces the estimated peak in hospital occupancy (i.e. VOC E+LH lies below VOC E in Figure 2b). In particular, given the full removal of NPIs from the outset of Step 4 (termed RM (roadmap) completion), our VOC E+LH scenario gave a mean peak occupancy below the January 2021 peak of 34,336 COVID-19 patients, whereas for VOC E the mean peak occupancy was approximately 60,000.

In addition to these four illustrative VOC characteristics (VOC MT, VOC E, VOC LT+E and VOC E+LH), additional sensitivity analyses of peak hospital occupancy to possible VOC efficacy and transmission are given in supplementary heat maps (Supplementary Fig. 12). We found that more severe immune escape and/or a variant with both immune escape and increased transmissibility would likely result in scenarios where reversion to more stringent NPI measures would be required to prevent hospitals being quickly overwhelmed.

The modelled outcomes involving large peaks in hospitalisations should be interpreted as being indicative of the relative extent of control measures required to keep the variant under control; we find that the resistance of the variant to current vaccines was the most significant indicator of how much measures may be safely relaxed. We stress that if there was a surge in hospital occupancy, shifts in public behaviour and enaction of national legislation may limit the spread of infection [51]. Therefore, our scenarios represent a pessimistic view of measures in response to a worsening outbreak.

We propose that the age-distribution of cases may give an early signal of whether a variant displays immune escape or higher transmissibility (Figure 2c). Previous infections to date have been higher in younger age groups who typically have higher rates of contact and are less likely to have been shielding to the same degree as more vulnerable age groups. As a result, with the relaxation of NPIs we might expect to see proportionally increased infection from resident variants in the older (60+ years) age groups. On the other hand, as vaccinations were largely offered first to older age groups, we might also expect to see a large proportional increase in infections amongst children (Figure 2c, grey bars vs black bars). Such effects were reduced for a VOC with increased transmission (e.g. VOC MT, blue bars), as it is expected to cause an earlier surge in cases at a time when the vaccination program is less advanced. VOCs with immune escape characteristics (VOC E, orange bars, and VOC LT+E, yellow bars) were less affected by both previous infection and vaccination, resulting in an age-distribution of infection that more closely matched the historical profile. Nonetheless, if the roadmap is run to completion, the more relaxed levels of NPIs than have been previously seen is still expected to cause significantly higher infections amongst the elderly than occurred to date.

### Early phase VOC dynamics and the implications of VOC-targeted vaccines

Our final piece of analysis explored the outbreak potential of putative VOCs and evaluated the impact on the infectious disease dynamics of the relative timing of a VOC-targeted vaccine, with improved efficacy towards VOCs, becoming available.

For a given transmissibility and level of effective imports per day (the daily rate of second generation cases that result from a single index case), we used the stochastic VOC importation model to calculate the epidemic probability, which we subjectively defined as the probability of reaching a prevalence of 100 cases within 365 days (Figure 3a). We discerned two prominent features. A variant that was less transmissible could be almost certain to become established if the effective importation rate was high enough. Epidemic probabilities of 1 were attained for relative transmissibilities of 0.7 (when effective imports per day were 0.22 or above), 0.8 (0.12 effective imports per day and above) and 0.9 (0.10 effective imports per day and above). This contrasts with a VOC that was substantially more transmissible than resident variants, where even at low numbers of effective imports per day (0.02 per day) it remained highly likely the VOC could become established; VOCs with a relative transmissibility of 1.3 or above returned epidemic probabilities above 0.9.

**Figure 3:**
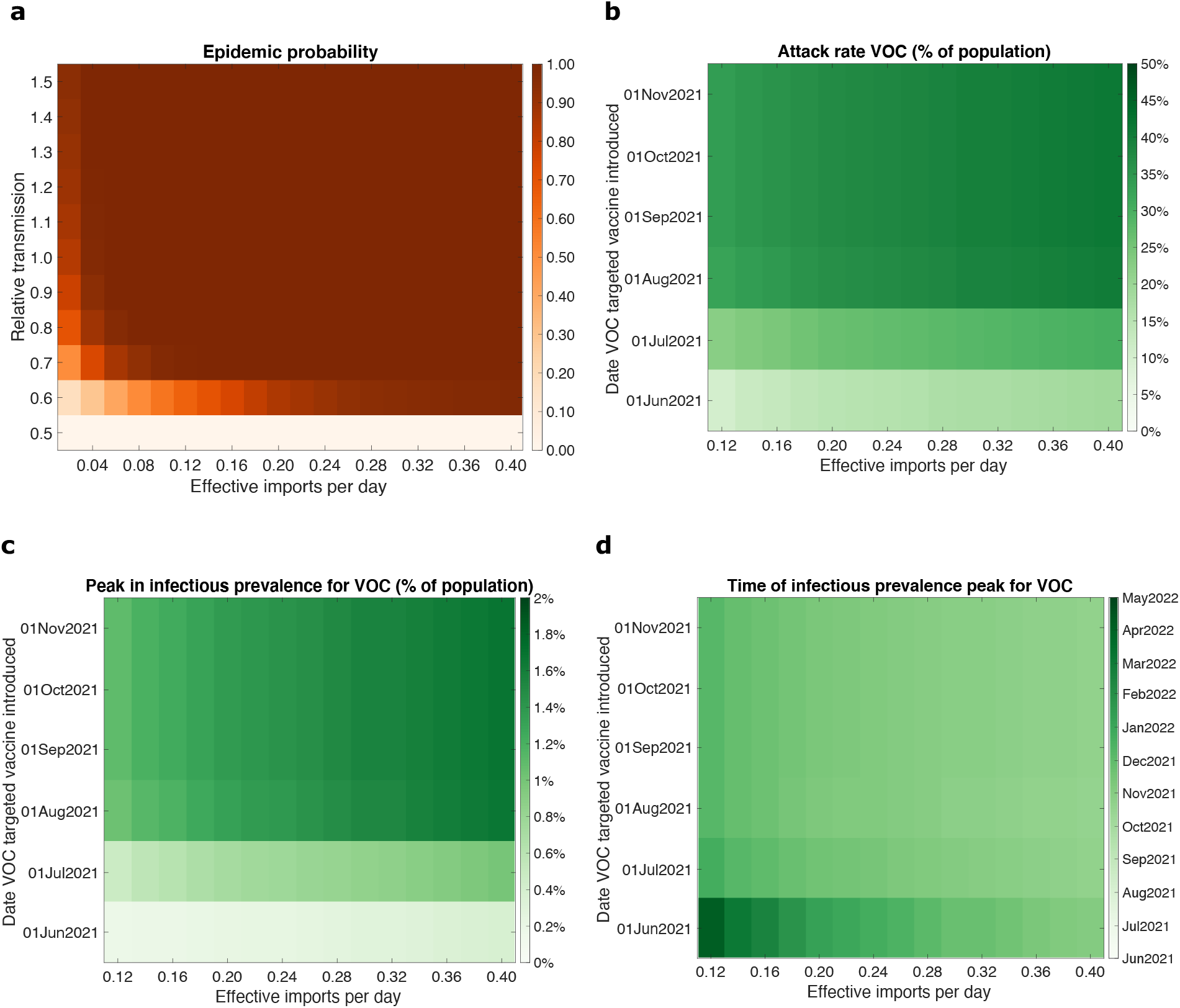
Outbreak potential and sensitivity of epidemic trajectories to the introduction time of a VOC targeted vaccine for VOC E. **a** The probability of an epidemic for varying relative transmissibilities (compared to resident variants) versus a given count of VOC effective imports per day (corresponding to the second generation cases that result from a single index case). In panels **b-d**, we performed simulations using the parsimonious SARS-CoV-2 transmission model for differing effective VOC importation counts and introduction date of a VOC targeted vaccine and evaluated the following epidemiological summary statistics for the resultant VOC outbreak: **b** final size; **c** peak in infectious prevalence; **d** time of peak in infectious prevalence.

Sampling from the stochastic VOC importation model to initialise the introduction time of 100 VOC infected individuals and their distribution across the applicable infected compartmental states, we next used the parsimonious SARS-CoV-2 transmission model to consider the sensitivity of the magnitude and timing of a VOC caused resurgence of SARS-CoV-2 infection. We found that the introduction date of a VOC targeted vaccine was much more important than the effective imports per day for the final size (Figure 3b), peak (Figure 3c) and time of peak (Figure 3d). Above all, if the VOC-targeted vaccine was not introduced until August 2021 or later, the VOC attack rate was close to 50% of the total population (Figure 3b), and the peak in infectious prevalence was in the region of 1-2% (Figure 3c) and occurred during September/October 2021 (Figure 3d).

Changing the prioritisation scheme for the VOC-targeted vaccine, to one in which unvaccinated individuals were given precedence followed by those who had received one of the pre-existing vaccines, resulted in qualitatively comparable findings (Supplementary Fig. 13).

## Discussion

Through a set of mathematical modelling analyses, we have demonstrated the epidemiological trajectories for putative VOCs to be wide-ranging and heavily dependent on its transmissibility and immune escape properties.

Our findings are in concordance with illustrative modelling of novel SARS-CoV-2 variants conducted in May 2021 by three academic groups in the UK contributing to SPI-M-O, which showed novel SARS-CoV-2 variants that either are highly transmissible or substantially escape immunity have the potential to lead to resurgences (in the absence of NPIs) in infections and hospitalisations that are larger than those seen in January 2021 in the UK [38]. Whilst the assumptions that lead to a resurgence in hospitalisations to levels comparable to those witnessed in the UK during January 2021 might seem extreme, SARS-CoV-2 has already demonstrated its adaptive potential. At the present time (July 2021), there is no reason to believe that the SARS-CoV-2 virus has yet settled at its fitness optimum in terms of replication and transmission capabilities. Given the prospect of the virus undergoing a continued accumulation of adaptive mutations, we should remain alert to all possible scenarios and continue an evidence-based analysis of evolutionary change so that public health measures can be adjusted in response to substantive changes in viral infectivity or severity of COVID-19 (also advocated by Day *et al*. [52]).

Our transmission modelling suggests that the ability of variants to evade immunity derived from vaccination (with currently available vaccines) can be a key indicator of how much measures may be relaxed without risking further surges of infection and cases requiring hospital care. Furthermore, alongside the relative size of peaks in infection and hospitalisation, their timing may also be of great importance. It is hoped that vaccines may by adapted to more effectively target emerging variants. Within our model framework and utilised assumptions, our work suggests a critical interplay between the timing of a VOC-targeted vaccine and the number of effective imports of a VOC. When the number of effective imports per day is sufficiently low (less than 0.5 per day in our model), it was possible for a new vaccine introduced early enough to have an appreciable effect on the VOC epidemic curve. With reports that the mass production of AZ vaccine requires 60 days to grow the cells followed by 28 days of quality assurance [53], one may reasonably expect an absolute minimum of three months from identification of a novel SARS-CoV-2 variant to the possible initial administration of revised vaccines that use the viral vector technology platform.

If a concerning novel variant is identified within a population, it is conceivable that the relationship between the prevalence of the variant and any change in NPI policy could give a signal of its characteristics. We would expect that a variant with no immune escape properties, but that is even more transmissible than resident variants, would display dynamics akin to the emergence and establishment of B.1.1.7 in the UK. In particular, over a period of fairly static NPIs, it was observed that whilst growth rates of resident variants were non-increasing, the B.1.1.7 variant had a positive growth rate [7, 8]. On the other hand, a variant with no transmission advantage, but displaying immune escape, could be identified through a shift in the distribution of cases between vaccinated and unvaccinated individuals. In addition, the timing of a surge in a novel variant could also give a clue as to its characteristics. As we continue with the vaccination rollout, reducing the level of NPIs could reveal less-transmissible immune-escape variants, which were previously kept in check by control measures. If sufficient data is available to track the distribution of variant cases between vaccinated and unvaccinated individuals, there is the potential to identify such a variant (such as the B.1.351 variant, for which reduced vaccine efficacy has previously been observed [22–26]) in advance of surging cases in response to reductions in measures, indicating the need for close surveillance as measures are lifted.

We suggest multiple courses of action that can act in concert to mitigate the risk of a widespread outbreak caused by a new VOC, summarised in the following four paragraphs comprising of: genomic surveillance; pharmaceutical interventions (therapeutics and vaccines); slowing rate of importations; and early detection efforts.

Genomic sequencing of SARS-CoV-2 viral samples is of paramount importance. A concerted international COVID-19 pandemic response requires global situational awareness of how the virus is mutating and identification of emergent variants that are of concern. The World Health Organization advocates strengthening surveillance and sequencing capacity, and a systematic approach to provide a representative indication of the extent of transmission of SARS-CoV-2 variants [1].

Together with support of research to develop treatments for mitigating disease impacts [54], it is crucial to maximise vaccine uptake and homogeneity in vaccine coverage to broaden immunity across the population. High SARS-CoV-2 incidence rates act to increase the vaccine escape risk, meaning keeping case numbers low using both NPIs and pharmaceutical measures is beneficial [2, 55]. Were vaccine escape variants to arise, one potential measure that could be taken would be a targeted booster vaccine against said variants. From a global standpoint, equitable vaccine distribution is also advocated, with it thought to decrease the potential for antigenic evolution [56].

Furthermore, there is reason to believe that slowing importation of new variants into the UK is an important priority to afford additional time to bolster vaccine-acquired immunity throughout the population, heighten surveillance procedures and build capacity for locally targeted interventions [57]. To that end, analysis of genomic and contact tracing data has demonstrated the efficacy of travel restriction policy (travel corridors) enacted in England over the summer of 2020 in reducing both the number of contacts reported by positive cases and the number of subsequent cases due to onward transmission [58].

Generally, any single cluster of infections with a VOC will be most easily controlled whilst the case count is small. Early detection and efforts to extinguish infection clusters is therefore paramount, as increased importation rates seed more clusters and will necessitate additional resources to keep a VOC under control. From 1st February 2021 in England, the government began using surge testing (in combination with genomic sequencing) in specific locations to monitor and suppress the spread of variants. At the original time of writing this manuscript (May 2021), surge testing involved increased testing, including of those without symptoms of COVID-19 and door-to-door testing in some areas, and enhanced tracing of close contacts of confirmed cases infected by the variant of concern [59].

Our model parameterisation, vaccine rollout and NPI policy were tailored to England; we would not expect our findings to be directly applicable in other countries and regions, though the broad messages may still be relevant. These results would transfer most readily to settings with a resident variant already in circulation, where NPIs of moderate stringency are in place and beginning to be relaxed in a phased manner, and vaccination is underway. In contrast, countries such as New Zealand and Australia that have not had large numbers of cases may have very different dynamics to the UK, and how they choose to control the development of population immunity will affect their response to new variants. The varying combinations of vaccines and how they are used in different countries will also affect how new variants may be discerned in the data, particularly in the age distribution of cases and hospitalisations. Nevertheless it is still likely to be the case that variants showing substantial vaccine escape may only become apparent once vaccine rollout is largely underway.

Our work demonstrates the use of parsimonious model structures to garner qualitative insights and high-level quantification of the order of magnitudes of public health measurable quantities of interest (such as hospitalisations and deaths) that may be experienced. Operationally, there is a balance between having a model of sufficient detail to provide robust insights on the objective and the time required to obtain such insight. Models with additional complexities typically require longer development times and finer-resolution data to be reliably parameterised. In addition, higher dimensional dynamical systems can result in parameter inference becoming more computationally intensive [60]. In a global public health emergency such as a pandemic, policy processes tend to be very fast. Using more limited methods to ensure the timely delivery of findings before a policy decision is taken can be worth more than using a more complex method and obtaining results afterwards, provided any methodological limitations are made clear [61]. That being said, incorporating noted heterogeneities in the infectious disease dynamics is a crucial consideration for interventions that are targeted according to those heterogeneities (such as the prioritisation order of COVID-19 vaccination in the UK being predominately determined by age).

Where possible, we have taken a data-driven approach to parameterise the models. Nevertheless, this work has made simplifying assumptions and our results therefore have limitations. We assumed no waning of immunity to a specific variant induced via natural infection or vaccination, and note that rapid waning of immunity would lead to more severe outcomes than presented here. Evidence suggests previous infection with SARS-CoV-2 induces effective immunity to future infections in most individuals, however natural protection for previously infected individuals can be temporary [62–65], although the robust quantification of reinfection risk is also complicated by variants. We also did not include any seasonal effects, that, if present, may impact the timing of future waves of infection [66]. Our analysis would also be affected by deviations from the vaccination programme included here, such as the rollout speed and split between the different vaccine types, or changes in NPI stringency (including the relaxation and/or strengthening of measures).

In summary, we have illustrated broad principles for the possible implications of the emergence of SARS-CoV-2 variants that have particular transmissibility and immune escape attributes. More transmissible or immune-escape variants may cause substantial waves of infection, even in the context of considerable vaccine-derived immunity. Indeed, a less-transmissible variant with (partial) immune escape could be revealed as NPIs are lifted, and cause an appreciable wave of infections, and even hospitalisations. The unpredictability in the epidemiological characteristics of novel pathogens mean our ideas and understanding can change as new information on the outbreak is accrued. Close monitoring of the evolution of SARS-CoV-2 across a range of geographical scales is needed to enhance local situational awareness and quantify risk from variants that may be of concern, with reliable and accurate data ensuring outputs from models of infectious disease dynamics are as informative as possible.

## Methods

We first overview the assumptions applied across all our models, then present in turn each model and the analyses that was performed in each case, before closing by summarising our vaccine efficacy assumptions.

### Model agnostic assumptions

Since we are primarily interested in the epidemiological impact of variants, in all models we assumed no waning immunity (for immunity resulting from either natural infection or vaccination), no ‘seasonality’ in the form of oscillatory rate constants, and no individual-level reinfection with the same variant. This allows our results to capture the pure signal from variant effects, although there is nothing in our approach that precludes inclusion of additional phenomena if they are of scientific or practical interest.

We assumed ‘leaky’ immunity in both our transmission models, so that when protective immunity acquired from natural infection and/or vaccination was imperfect (0 *< ϵ <* 1, with 0 corresponding to no protection and 1 complete protection), individuals experience a reduced, but non-zero risk (i.e. susceptibility of 1 *− ϵ*). The ‘leaky’ mechanism contrasts with an ‘all-or-nothing’ immunity conceptualisation, where a proportion *E* of the population would be fully protected and the remaining proportion (1 *− ϵ*) are fully unprotected. For the portion of the vaccinated population that had been previously infected by a variant, we set the level of protective immunity at the greater amount of immunity attained between the two types of exposure. We also assumed that for those previously infected by either the resident variant or the VOC, the prior-infection efficacies towards the other variant were identical (i.e. the level of protection towards the VOC given a prior resident variant infection matched the level of protection towards the resident variant given a prior VOC infection).

Both transmission models introduced 2,000 VOC infected individuals (a prevalence of approximately 0.0035%) on 17th May 2021 unless stated otherwise, representing a comparable population to the new non-B.1.1.7 VOCs reported in England in early to mid-May 2021 [36]. We modelled the co-circulation of the VOC and resident variant (for further details, see the model descriptions below and Sections 2 and 3 of the Supplementary Information). We consider the initial group of VOC infected individuals to be large enough that the average dynamics are reasonably captured by a deterministic system (see Section 4.4 of the Supplementary Information). We took the number of initial VOC infected individuals from the portion of the population that were both unvaccinated and not previously infected by any variant, with the VOC then considered in co-circulation with the resident variant.

To capture changes in contact/mobility in response to relaxations of NPIs at each Step of the roadmap out of lockdown for England [37], we set estimates of R excluding immunity in each Step at central values used by the University of Warwick SARS-CoV-2 transmission model for modelling work assessing the relaxation of restrictions (Roadmap Step 3 modelling [38]). For the breakdown of R excluding immunity values within each step, and the associated time intervals, see Table 3.

The models included vaccinations with AZ, Pfizer and Moderna vaccines, with the latter two considered equivalent. We assumed a vaccine rollout speed averaging 2.7 million doses per week until the week commencing 19th July 2021 and 2 million doses per week thereafter (based on the central roll-out speed scenario provided by Cabinet Office to the Scientific Pandemic Influenza group on Modelling, Operational subgroup (SPI-M-O) for use in modelling of easing restrictions: Roadmap Step 3 [38]).

We performed all model computations using Matlab R2021a.

### The parsimonious SARS-CoV-2 transmission model

#### Model description

We developed a parsimonious deterministic ordinary differential equation (ODE) model consisting of an SEIR disease state formulation for resident variants (including B.1.1.7) and a VOC (see Table 3 for parameterisation), with variant-specific transmissibility. Model equations can be found in Section 2 of the Supplementary Information.

We initialised the proportion of the population vaccinated with the AZ vaccine and Pfizer/Moderna vaccines using data reported from the National Immunisation Management Service (NIMS), the System of Record for the NHS COVID-19 vaccination programme in England [39] (see Table 3). We used the vaccine rollout speed to calculate the number of first doses administered per day. We assumed a future mix of vaccinations in the ratio 60% (AZ), 30% (Pfizer) and 10% (Moderna) (as used in [40]). Where individuals were both recovered and vaccinated, we assumed they received the greater of the two protections.

We used a population size of 56 million, comparable to the ONS mid-2019 estimate for the population of England [41]. All simulations began from 17th May 2021 with a time horizon of 365 days.

#### Investigating VOC outbreak potential and epidemic trajectories

We investigated the potential for illustrative VOCs to transmit widely amongst the community upon its introduction by computing the value of ‘R with immunity’ over time (also referred to as effective R), with notation 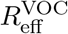. R with immunity includes both the effects of immunity (due to vaccination or prior infection) and the level of NPIs in place at the time. Thus, R with immunity represents the potential for a newly-introduced VOC to generate a large epidemic at a particular time, assuming that there is no deviation from the roadmap prior to that time.

#### Exploration of parameter sensitivity

We next explored how varying the transmissibility and immune escape attributes affected the outbreak size, peak in any resultant wave of infection and R with immunity. Our sensitivity used a range of relative transmissibilities of the VOC versus the resident variants (from 0.5 to 1.5, with an increment of 0.1), and a proportional efficacy against the VOC derived from vaccination or prior natural-infection (from 0.5 to 1.0, with an increment of 0.05).

### The age-structured SARS-CoV-2 transmission model

#### Model description

In the previously outlined parsimonious model, we did not include age-structure and only considered SARS-CoV-2 infections. To assess the healthcare implications of a VOC that becomes established amongst the community, we extended the University of Warwick SEIR-type compartmental age-structured model, developed to simulate the spread of SARS-CoV-2 within regions of the UK [42], to allow inclusion of a putative VOC.

The model has been fitted to UK outbreak data, giving a comparable match to deaths, hospital admissions, hospital occupancy and test positivity from community testing (Pillar 2 tests). The model is formulated as a system of ordinary differential equations (Section 3 of the Supplementary Information).

The force of infection for this model was determined by the use of age-dependent (who acquires infection from whom) social contact matrices for the UK [44, 45]. We assumed susceptibility and the probabilities of becoming symptomatic, being hospitalised and mortality to be age-dependent. Our model formulation accounted for the role of household isolation by allowing first infections within a household to cause new secondary infections at an increased rate (more details may be found in Keeling *et al*. [42]). This model construction allows secondary household contacts to be isolated and consequently play no further role in the outbreak.

#### Sensitivity of hospitalisations to VOC characteristics

Echoing the observed behaviour of COVID-19 infections, our age-structured SARS-CoV-2 transmission model differentiates between individuals who are symptomatic and those who are asymptomatic. Partitioning those infectious by symptom status allows for the lower level of transmission believed to be associated with asymptomatic infection. It also generates the possible progression of symptoms increasing in severity, leading to hospitalisation and/or death. For additional details on the calculation of hospital admissions, hospital occupancy and deaths from the number of new symptomatic infections on a given day, see Keeling *et al*. [42].

Utilising the case severity module of the model, we investigated daily hospital admissions, total hospital admissions and the impact on the infection age-distribution for our four main VOC scenarios: VOC MT, VOC E, VOC E+LH and VOC LT+E (Table 1). We simulated each VOC alongside the existing resident variants by the duplication of the base model equations (Section 3 of the Supplementary Information).

All simulations began from January 2020 (coinciding with the time the SARS-CoV-2 virus was first identified in England) with a time horizon of 1095 days (through to December 2022), although results presented here are abridged due to uncertainties arising from a rapidly evolving epidemic. The central estimates for *R* excluding immunity for Steps 3 and 4 matched those listed in Table 3.

### VOC outbreak potential and utility of VOC targeted vaccines

The previous analyses sought to evaluate the likely timescales, drivers and healthcare impact of SARS-CoV-2 VOC epidemics under a specific set of assumptions, along with their sensitivity to the variation in epidemiological parameters that underpin the transmission dynamics.

However, epidemics starting from a small number of seed introductions are inherently stochastic, and deterministic models are unable to capture that stochasticity. To that end, we adopted a stochastic modelling approach to explore the outbreak potential of a VOC post-emergence.

Our VOC importation model was a Gillespie stochastic simulation [46, 47] with six types-at-birth and 12 disease states. The types-at-birth comprised combinatorial combinations of two infection history states (either having had no prior-infection or to have been previously infected with resident variants) and three vaccination states (unvaccinated, vaccinated with AZ, vaccinated with Pfizer/Moderna). Infected individuals in these types could then be either latent infected or infectious, resulting in a total of 12 disease states. Using the stochastic framework, we studied the dependence of the epidemic probability (the probability of reaching a prevalence of 100 cases within 365 days) for a VOC E type variant on its relative transmissibility compared to resident variants (from 0.5 to 1.5, with an increment of 0.1) and on the amount of effective importations per day (from 0.02 to 0.40, with an increment of 0.02). We interpret importations as the second generation cases stemming from onward transmission to contacts of a single index case. To cross-check the correctness of the simulation, we compared it with analytical results for a continuous-time multi-type branching process model with immigration (see Section 4 of the Supplementary Information).

Furthermore, another uncertain aspect of the system is the plethora of SARS-CoV-2 vaccines in development [48] and the prospect of previously approved vaccine formulations being updated to improve protection against VOCs. For example, there has been in-vivo evidence regarding the efficacy of the Novavax vaccine against the B.1.351 variant from phase 2 trials in South Africa, finding 51.0% (95% CI: -0.6% to 76.2%) mild to moderate disease efficacy against B.1.351 in HIV negative individuals [49].

Using the parsimonious SARS-CoV-2 two-variant transmission model, we investigated the sensitivity of a VOC with immune escape properties (VOC E) to the timing and properties of a VOC-targeted vaccine. We sampled from the stochastic VOC importation model to initialise the introduction time of 100 VOC infecteds and their distribution across the applicable infected compartmental states of the parsimonious SARS-CoV-2 transmission model. We then appraised sensitivity to the date a VOC targeted vaccine began to be administered (from 1st June 2021 to 1st November 2021, with an increment of one month) versus the amount of effective VOC imports per day (from 0.12 to 0.40, with an increment of 0.02).

We assumed individuals previously vaccinated were subsequently re-vaccinated, exploring prioritisation to receive the VOC-targeted vaccine being either initially given to previously vaccinated individuals or to unvaccinated individuals. In all the above-described scenarios we fixed the R excluding immunity for resident variants in the stochastic model at 3.

### Vaccine efficacy assumptions

#### Mechanisms of vaccine action

The protective actions of vaccination can be separated into five components: (i) efficacy against infection; (ii) efficacy against symptomatic disease; (iii) efficacy against hospital admission; (iv) efficacy against death and (v) efficacy against onward transmission. Three vaccines are now in use in the UK (Pfizer, AstraZeneca and Moderna). As vaccine efficacies for Moderna are not currently as well defined from population-level observations (at the original time of writing in May 2021), we assumed equal efficacies for both Moderna and Pfizer vaccines, since they are both mRNA vaccines.

For the parsimonious deterministic model, as part of the parsimonious approach, we assumed the total vaccine infection efficacy effect was obtained after a single dose and that there was no delay in the onset of protective effects post-vaccination. We did not use efficacy estimates for symptomatic disease, hospitalisation or death as the parsimonious model tracked infections only.

In the age-structured SARS-CoV-2 transmission model, the effect of vaccination was realised at each stage of case severity progression, including parameters, with increases for each between one and two doses, for: (i) reduced infection, (ii) reduced symptoms, (iii) reduced hospitalisations (severe case outcomes). As well as corresponding to protection of the individual, symptom efficacy also had an impact on disease spread due to the model assumption that asymptomatic infected individuals, compared to symptomatic infected individuals, transmitted the virus at a reduced rate (i.e. were less infectious). Another assumption was that prevention of symptoms may be less affected by immune escape than infection; we fixed symptom efficacy at 90% of the estimated efficacy against resident variants for VOC E (compared to 75% for infection efficacy). Symptom efficacy also provided a lower bound for efficacy against hospitalisation, the latter taken between a 10% and 25% reduction for VOC E. We present full details of all efficacies used in Table 2, including the protection realised by previous infection in each of the three actions considered by vaccination. We assumed previously infected individuals to have equal protection regardless of vaccination status and we carried out a sensitivity analysis to explore a broader range of efficacy effects. We also used the age-structured model to assess the impact on hospitalisations, considering a VOC with similar characteristics to VOC E except with proportional efficacy against severe disease (hospitalisations) being unadjusted. We labelled this scenario as VOC E+LH (immune escape plus less hospitalisations).

**Table 2:** Efficacy assumptions against the resident variants and for our illustrative VOCs with immune ecscape (VOC E and VOC LT+E, with efficacies for these VOCs stated in parentheses). We computed the efficacies against VOCs as the product of the proportional vaccine efficacy and the efficacy against the resident variants. For a summary table of source studies, see Supplementary Table 1.

|  |  | Pfizer |  | AstraZeneca |  | Natural immunity |
| --- | --- | --- | --- | --- | --- | --- |
|  | Efficacy action | Dose 1 | Dose 2 | Dose 1 | Dose 2 |  |
| **Parsimonious model** | **Infection** | 75% (56%) |  | 65% (49%) |  | 100% (75%) |
| **Age-structured model** | **Infection** | 60% (45%) | 85% (64%) | 60% (45%) | 65% (49%) | 100% (75%) |
|  | **Symptoms** | 60% (54%) | 90% (81%) | 60% (54%) | 80% (72%) | 100% (90%) |
|  | **Hospitalisation** | 80% (60%) | 90% (81%) | 80% (60%) | 90% (72%) | 100% (90%) |

**Table 3:**
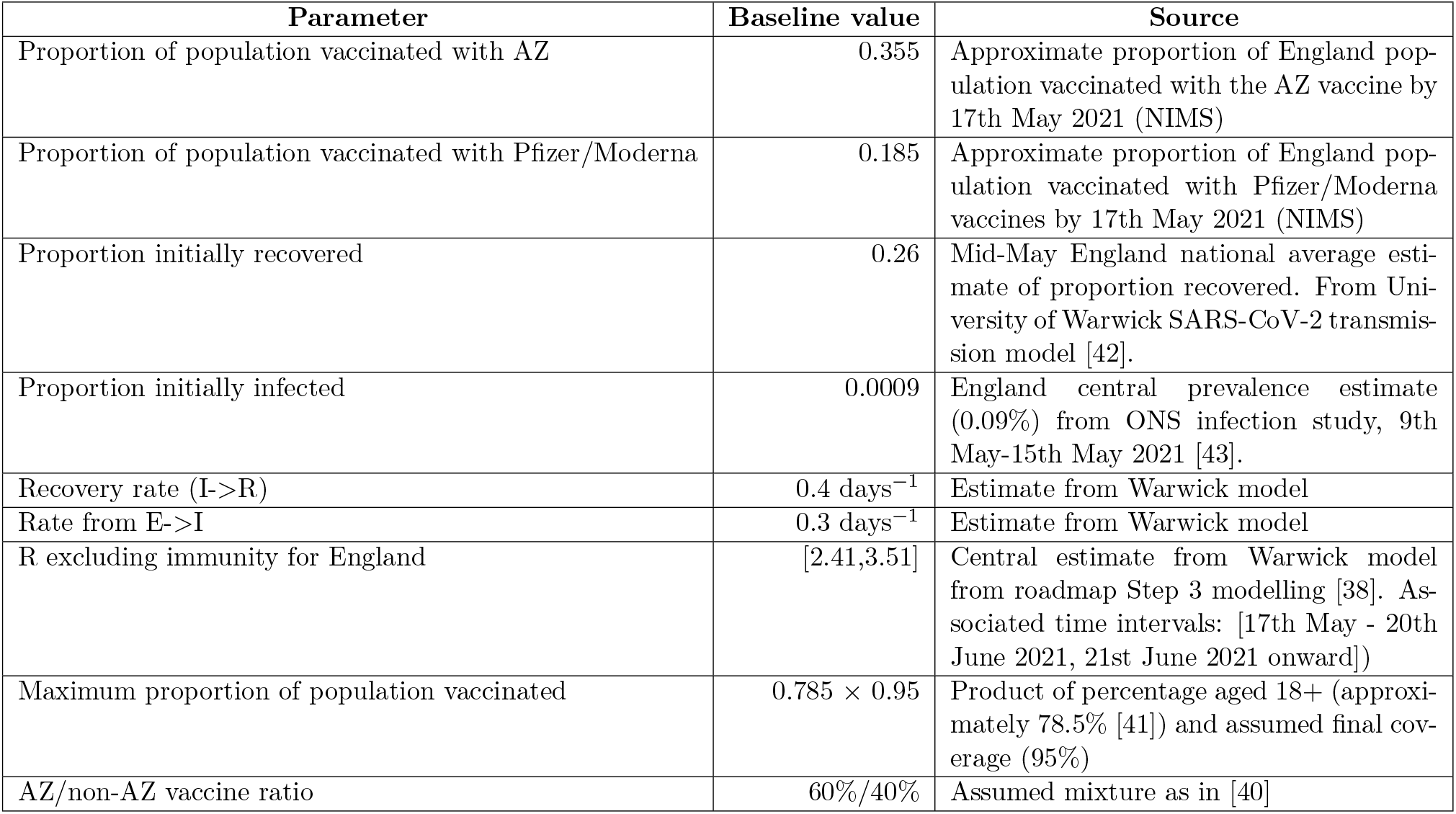
Initial conditions, epidemiological and vaccination parameter assumptions for the parsimonious SARS-CoV-2 transmission model.

| Parameter | Baseline value | Source |
| --- | --- | --- |
| Proportion of population vaccinated with AZ | 0.355 | Approximate proportion of England population vaccinated with the AZ vaccine by 17th May 2021 (NIMS) |
| Proportion of population vaccinated with Pfizer/Moderna | 0.185 | Approximate proportion of England population vaccinated with Pfizer/Moderna vaccines by 17th May 2021 (NIMS) |
| Proportion initially recovered | 0.26 | Mid-May England national average estimate of proportion recovered. From University of Warwick SARS-CoV-2 transmission model [42]. |
| Proportion initially infected | 0.0009 | England central prevalence estimate (0.09%) from ONS infection study, 9th May-15th May 2021 [43]. |
| Recovery rate ($I \rightarrow R$) | $0.4 \text{ days}^{-1}$ | Estimate from Warwick model |
| Rate from $E \rightarrow I$ | $0.3 \text{ days}^{-1}$ | Estimate from Warwick model |
| R excluding immunity for England | [2.41, 3.51] | Central estimate from Warwick model from roadmap Step 3 modelling [38]. Associated time intervals: [17th May - 20th June 2021, 21st June 2021 onward]) |
| Maximum proportion of population vaccinated | $0.785 \times 0.95$ | Product of percentage aged 18+ (approximately 78.5% [41]) and assumed final coverage (95%) |
| AZ/non-AZ vaccine ratio | 60%/40% | Assumed mixture as in [40] |

#### Vaccine efficacy estimates against resident variants

Central vaccine efficacy estimates for both transmission models (Table 2) are based on the emerging data in the UK population and elsewhere. Source studies for these estimates can be found in Supplementary Table 1.

#### Vaccine efficacy estimates against VOCs

As of May 2021, there was limited evidence (though ever increasing) regarding the efficacy of the various vaccines against VOCs, such as B.1.351 and B.1.617.2, and the susceptibility of individuals with prior infection by resident variants (including the B.1.1.7 variant) to other VOCs. Available estimates (as of May 2021) from the literature on the efficacy of vaccine-induced and naturally-acquired immunity can be found in the Supplementary Information, Sections 1.2-1.3.

We summarise the transmissibility and infection immune escape properties for each of our putative VOCs in Table 1 as proportions compared to the resident strains, with the efficacies for a subset of our illustrative VOCs (VOC E and VOC LT+E) provided in Table 2.

## Data Availability

The data utilised in this study are publicly available, with relevant references and data repositories stated within the main manuscript and Supporting Information. The code repository for the study is available at: https://github.com/LouiseDyson/COVID19-variants-of-concern-modelling-paper.

## Acknowledgements

We are grateful to Public Health England for providing data from the National Immunisation Management Service (NIMS), the System of Record for the NHS COVID-19 vaccination programme in England. We thank the members of the JUNIPER consortium for helpful comments on the manuscript.

SM and MJK were supported by the National Institute for Health Research (NIHR) [Policy Research Programme, Mathematical & Economic Modelling for Vaccination and Immunisation Evaluation, and Emergency Response; NIHR200411]. The views expressed are those of the authors and not necessarily those of the NIHR or the Department of Health and Social Care. LD, EMH, MJT and MJK were supported by the Medical Research Council through the COVID-19 Rapid Response Rolling Call [grant number MR/V009761/1]; LD, MJT and MJK were supported by the Engineering and Physical Sciences Research Council through the MathSys CDT [grant number EP/S022244/1]; KAL was supported by the Li Ka Shing Foundation; LP was supported by the Wellcome Trust and the Royal Society [grant number 202562/Z/16/Z]; TH was supported by the Royal Society [grant number INF/R2/180067]; TH and LP were also supported by the UK Research and Innovation COVID-19 rolling scheme [grant numbers EP/V027468/1 and MR/V028618/1] as well as the Alan Turing Institute for Data Science and Artificial Intelligence. LD, LP, TH, MJT and MJK were supported by UKRI through the JUNIPER modelling consortium [grant number MR/V038613/1]. The funders had no role in study design, data collection and analysis, decision to publish, or preparation of the manuscript.

## Author contributions

**Louise Dyson:** Conceptualisation, Data curation, Formal analysis, Funding acquisition, Methodology, Software, Supervision, Validation, Visualisation, Writing - Original draft, Writing - Review & Editing.

**Edward M. Hill:** Conceptualisation, Data curation, Formal analysis, Methodology, Software, Validation, Visualisation, Writing - Original draft, Writing - Review & Editing.

**Sam Moore:** Data curation, Formal analysis, Methodology, Software, Validation, Visualisation, Writing - Original draft, Writing - Review & Editing.

**Jacob Curran-Sebastian:** Formal analysis, Methodology, Software, Validation, Visualisation, Writing - Review & Editing.

**Michael J. Tildesley:** Conceptualisation, Funding acquisition, Methodology, Supervision, Writing - Review & Editing.

**Katrina A Lythgoe:** Conceptualisation, Writing - Review & Editing.

**Thomas House:** Conceptualisation, Formal analysis, Funding acquisition, Methodology, Software, Supervision, Validation, Visualisation, Writing - Review & Editing.

**Lorenzo Pellis:** Conceptualisation, Formal analysis, Funding acquisition, Methodology, Software, Supervision, Validation, Visualisation, Writing - Review & Editing.

**Matt J. Keeling:** Conceptualisation, Funding acquisition, Methodology, Supervision, Writing - Review & Editing.

## Competing interests

All authors declare that they have no competing interests.

### Data availability

The data used in this study are provided in the GitHub repository associated with the study: https://github.com/LouiseDyson/COVID19-variants-of-concern-modelling-paper[67].

## Code availability

The code repository for the study is available at: https://github.com/LouiseDyson/COVID19-variants-of-concern-modelling-paper [67].

