## Supplementary Information for "Possible future waves of SARS-CoV-2 infection generated by variants of concern with a range of characteristics"

### 1 Summary of immunity efficacy estimates from the literature

We overview available estimates from the literature on efficacy of vaccine-induced and naturally-acquired immunity as of May 2021.

#### 1.1 Efficacy of vaccine-induced immunity against resident variants

We collate in Supplementary Table 1 estimates of vaccine efficacy against the resident variants in the UK (during the time period when the B.1.1.7 variant was predominant), gathered across clinical trial data and through real-world observational studies of vaccination impact using surveillance data.

**Supplementary Table 1:** Vaccine efficacies with citations (A-K) and structured-model values (bold). Citations are: A. SIREN study data [1]. B. Data from Clalit Health Services, the largest of four integrated health care organisations in Israel, which insures 4.7 million patients (53% of the population) [2]. C. Nationwide surveillance data from Israel between 24th January-3rd April 2021 [3]. D. Phase 3 trial data for the AZ vaccine [4]. E. PHE analysis of trial data. F. PHE analysis using a test negative case control to estimate vaccine effectiveness against symptomatic disease [5]. G. Phase 3 trial data for the Pfizer-BioNTech vaccine [6]. H. Public Health Scotland analysis [7]. I. PHE analysis of Pillar 2 community test data on efficacy against hospitalisation among older adults in England [8]. J. Bristol Hospital analysis [9]. K. HOSTED study [10].

| | **Pfizer** | | **AZ** | |
|---|---|---|---|---|
| | **1st Dose** | **2nd Dose** | **1st Dose** | **2nd Dose** |
| **Efficacy against infection** | 70% (55-85%)$^{A}$<br>46% (40-51%)$^{B}$<br>**60%** | 85% (74-96%)$^{A}$<br>92% (88-95%)$^{B}$<br>95% (95-96%)$^{C}$<br>**85%** | 64% (46-77%)$^{D}$<br>**60%** | 60% (36-75%)$^{D}$<br>**65%** |
| **Efficacy against symptoms** | 57% (50-63%)$^{B}$<br>91% (74-97%)$^{E}$<br>49% (43-55%)$^{F}$<br>52% (29-69%)$^{G}$<br>**60%** | 94% (87-98%)$^{B}$<br>97% (97-97%)$^{C}$<br>93% (90-96%)$^{F}$<br>95% (90-98%)$^{G}$<br>95% (90-98%)$^{H}$<br>**90%** | 76% (59-86%)$^{D}$<br>51% (47-55%)$^{F}$<br>**60%** | 81% (60-91%)$^{D}$<br>66% (54-75%)$^{F}$<br>**80%** |
| **Efficacy against hospital admissions** | 74% (46-86%)$^{B}$<br>85% (76-91%)$^{H}$<br>73% (60-81%)$^{I(80+)}$<br>84% (74-89%)$^{I(70-79)}$<br>79% (47-92%)$^{J}$<br>**80%** | 87% (55-100%)$^{B}$<br>97% (97-97%)$^{C}$<br>**90%** | 94% (73-99%)$^{H}$<br>81% (76-85%)$^{I(80+)}$<br>81% (73-87%)$^{I(70-79)}$<br>80% (36-94%)$^{J}$<br>**80%** | 93% (89-95%)%$^{I(80+)}$<br>**90%** |
| **Efficacy against onward transmission** | 49% (38-58%)$^{K}$<br>**50%** | **50%** | 38% (21-52%)$^{K}$<br>**40%** | **50%** |

#### 1.2 Efficacy of vaccine-induced immunity against VOCs

The evidence base informing the vaccine efficacy offered by the presently available Pfizer-BioNTech and AZ vaccines against VOCs is expanding. We summarise vaccine efficacy estimates against the B.1.351 and B.1.617.2 variants, with estimates compiled in May 2021.

**Efficacy of vaccine-induced immunity against B.1.351**

From clinical trial data for the AZ vaccine, one study found the vaccine to have an efficacy of 10.4% (95% CI: -76.8% to 54.8%) against mild to moderate disease caused by the B.1.351 variant [11].

There is the prospect of more data on vaccine efficacy against the B.1.351 variant being forthcoming from surveillance data in nations that have made the greatest progress to date with vaccination rollout. For example, in Israel, there has been increased breakthrough rates of SARS-CoV-2 variants of concern in Pfizer-BioNTech vaccinated individuals, including B.1.351 [12]). In Qatar, Pfizer-BioNTech vaccine effectiveness estimates against severe, critical, or fatal disease due to infection with any SARS-CoV-2 virus (with the B.1.1.7 and B.1.351 variants being predominant within Qatar), were very high, at 97.4% (95% CI: 92.2 to 99.5) [13].

We may also draw insights from neutralisation studies. Prior work modelling the relationship between in vitro neutralisation levels and observed protection from SARS-CoV-2 infection has shown neutralisation levels to be predictive of immune protection [14]. That being said, sample sizes in neutralisation studies have been small, meaning there is considerable uncertainty. We also recognise that, though neutralising antibodies serve as one line of defense against infection, the T cell response is also important for prevention of further disease progression [15].

For the Pfizer-BioNTech vaccine, neutralization titers were reduced 8- to 9-fold against B.1.351 [16]. Planas *et al.* [17] found that three weeks after the first dose , 38% and 0% of serum samples neutralized B.1.1.7 and B.1.351 strains, respectively, compared to 92% (B.1.1.7) and 77% (B.1.351, with a low titer) three weeks after the second dose (sera from 16 vaccine recipients).

**Efficacy of vaccine-induced immunity against B.1.617.2**

An observational study using surveillance data in England aimed to estimate the effectiveness of the AZ and Pfizer vaccines against symptomatic disease with the B.1.617.2 variant. The analysis included data for all age groups from 5th April 2021 to cover the period since the B.1.617.2 variant emerged and it included 1,054 people confirmed as having the B.1.617.2 variant through genomic sequencing, including participants of several ethnicities. The study found that, for the period from 5th April to 16th May 2021, vaccine effectiveness against symptomatic disease from the B.1.617.2 variant was similar after two doses compared to the B.1.1.7 variant dominant in the UK. It is also expected to see even higher levels of effectiveness against hospitalisation and death [5].

In further detail, the Pfizer-BioNTech vaccine was 87.9% effective (95% CI: 78.2% to 93.2%) against symptomatic disease from the B.1.617.2 variant 2 weeks after the second dose. Two doses of the AstraZeneca vaccine were 59.8% effective (95% CI: 28.9% to 77.3%) against symptomatic disease from the B.1.617.2 variant.

Both vaccines were found to be approximately 33% effective against symptomatic disease from B.1.617.2, three weeks after the first dose; the first dose effectiveness estimate for the Pfizer vaccine was 33.2% (95% CI: 8.3% to 51.4%) and for the AZ vaccine was 32.9% (95% CI: 19.3% to 44.3%).

#### 1.3 Efficacy of naturally-acquired immunity against VOCs

A neutralisation study by Planas *et al.* [17] observed a loss of activity against B.1.351 in approximately 40% of convalescent sera after 9 months (83 sera from 58 individuals). We note that though these sera are not necessarily from those previously infected with B.1.1.7 specifically, the authors did observe neutralisation against B.1.1.7 in approximately 95% of these sera up to 9 months.

There is support for recovered individuals from each variant experiencing a (potentially asymmetric) degree of cross-immunity. A comparison of sera from infection with B.1.351 or the parental variant

B.1.1.117 in South Africa observed that B.1.351 infection induced substantial cross-neutralisation of the parental variant B.1.1.117 [18]. In contrast, antibodies elicited by B.1.1.7 infection have been shown to be less cross-reactive with other dominant SARS-CoV-2 strains than those induced by the parental strain [19].

#### 2 Parsimonious SARS-CoV-2 transmission model description

The model consists of an SEIR system for two variants, and also includes vaccination status. We follow the populations given by $N_{i,j,v}$, where $i$ and $j$ denote whether the population is susceptible ($S$), exposed ($E$), infectious ($I$) or recovered ($R$) to resident variants and VOC, respectively, and $v$ denotes vaccination status as unvaccinated ($u$), or vaccinated with the AstraZeneca vaccine ($a$), Pfizer-BioNTech or Moderna vaccines ($p$), or an updated/new vaccine ($n$).

For convenience, we define the effective number of infectious individuals with the resident variants in the UK ($I_{\text{UK}}$) or VOC ($I_{\text{VOC}}$) as

$$I_{\text{UK}} = \sum_{j,v} t_{I,j,v} N_{I,j,v}, \qquad I_{\text{VOC}} = \sum_{i,v} t_{i,I,v} N_{i,I,v},$$

where $t_{I,j,v} \in [0,1]$ and $t_{i,I,v} \in [0,1]$ corresponds to the transmissibility of infection for the resident variants infectious population compartment and VOC infectious population compartment, respectively; a value of 1 corresponds to no adjustment in transmission due to vaccination or prior infection, whereas values less than 1 correspond to a reduction in transmission (a transmission blocking effect) due to vaccination and/or prior infection. For example, $t_{I,R,u} = 0.5$ corresponds to a relative transmissibility of 0.5 (50% reduction in transmissibility) of the resident variants for those previously infected by the VOC (i.e is in the recovered state for the VOC) and who are unvaccinated. As another example, $t_{S,I,a} = 0.7$ corresponds to a relative transmissibility of 0.7 (30% reduction in transmissibility) of the VOC for those with no prior infection (i.e still susceptible to the resident variants) and who are vaccinated with the AZ vaccine.

Unless stated otherwise, we set $t_{I,j,v} = t_{i,I,v} = 1$ for all groups. For our simulations assessing sensitivity to a possible transmission blocking action of immunity, we set $t_{I,S,\{a,p,n\}} = t_{S,I,\{a,p,n\}} = t_{I,R,v} = t_{R,I,v} = \hat{t}$, with $\hat{t} \in [0,1]$ the modified transmissibility (equivalent to a 100(1-$\hat{t}$)% transmission blocking effect).

The model equations are given, for the unvaccinated, by:

$$\begin{aligned}
\frac{dN_{S,S,u}}{dt} &= -(\beta_{\text{UK}} I_{\text{UK}} + \beta_{\text{VOC}} I_{\text{VOC}} + V_A + V_P + V_n) N_{S,S,u}, \\
\frac{dN_{E,S,u}}{dt} &= (\beta_{\text{UK}} I_{\text{UK}}) N_{S,S,u}, \\
\frac{dN_{S,E,u}}{dt} &= (\beta_{\text{VOC}} I_{\text{VOC}}) N_{S,S,u}, \\
\frac{dN_{S,R,u}}{dt} &= -(\beta_{\text{UK}} s_u \text{UK} I_{\text{UK}} + V_A + V_P + V_n) N_{S,R,u}, \\
\frac{dN_{E,R,u}}{dt} &= (\beta_{\text{UK}} s_u \text{UK} I_{\text{UK}}) N_{S,R,u}, \\
\frac{dN_{R,S,u}}{dt} &= -(\beta_{\text{VOC}} s_u \text{VOC} I_{\text{VOC}} + V_A + V_P + V_n) N_{R,S,u}, \\
\frac{dN_{R,E,u}}{dt} &= (\beta_{\text{VOC}} s_u \text{VOC} I_{\text{VOC}}) N_{R,S,u}, \\
\frac{dN_{R,R,u}}{dt} &= -(V_A + V_P + V_n) N_{R,R,u},
\end{aligned} \tag{1}$$

where: $\beta_{\mathrm{UK}}$ and $\beta_{\mathrm{VOC}}$ are the transmission rates of the resident variants and VOCs, respectively; $s_{u\mathrm{UK}}$ is the susceptibility of unvaccinated VOC recovereds against infection with resident variants; $s_{u\mathrm{VOC}}$ is the susceptibility of unvaccinated resident variant recovereds against infection with the VOC; and $V_v$ represents vaccination with vaccine $v$. The model equations for the vaccinated are:

$$\begin{aligned}
\frac{dN_{S,S,v}}{dt} &= V_v N_{S,S,u} - (\beta_{\mathrm{UK}} e_{v\mathrm{UK}} I_{\mathrm{UK}} + \beta_{\mathrm{VOC}} e_{v\mathrm{VOC}} I_{\mathrm{VOC}}) N_{S,S,v}, \\
\frac{dN_{E,S,v}}{dt} &= (\beta_{\mathrm{UK}} e_{v\mathrm{UK}} I_{\mathrm{UK}}) N_{S,S,v}, \\
\frac{dN_{S,E,v}}{dt} &= (\beta_{\mathrm{VOC}} e_{v\mathrm{VOC}} I_{\mathrm{VOC}}) N_{S,S,v}, \\
\frac{dN_{S,R,v}}{dt} &= V_v N_{S,R,u} - (\beta_{\mathrm{UK}} s_{v\mathrm{UK}} I_{\mathrm{UK}}) N_{S,R,v}, \\
\frac{dN_{E,R,v}}{dt} &= (\beta_{\mathrm{UK}} s_{v\mathrm{UK}} I_{\mathrm{UK}}) N_{S,R,v}, \\
\frac{dN_{R,S,v}}{dt} &= V_v N_{R,S,u} - (\beta_{\mathrm{VOC}} s_{v\mathrm{VOC}} I_{\mathrm{VOC}}) N_{R,S,v}, \\
\frac{dN_{R,E,v}}{dt} &= (\beta_{\mathrm{VOC}} s_{v\mathrm{VOC}} I_{\mathrm{VOC}}) N_{R,S,v}, \\
\frac{dN_{R,R,v}}{dt} &= V_v N_{R,R,u}
\end{aligned} \tag{2}$$

where: $v \in \{a, p, n\}$; $e_{v\mathrm{UK}}$ and $e_{v\mathrm{VOC}}$ give the efficacy of vaccine status $v$ against resident variants and VOCs, respectively; and $s_{v\mathrm{UK}}$ and $s_{v\mathrm{VOC}}$ gives the susceptibility of VOC (or resident variants, respectively) recovereds with vaccination status $v$ against the resident variants (or VOC, respectively). Where individuals are both recovered and vaccinated, we assume they receive the greater of the two immune protections.

The disease progression equations are given by:

$$\begin{aligned}
\frac{dN_{E,j,v}}{dt} &= -\alpha N_{E,j,v}, & \frac{dN_{I,j,v}}{dt} &= -\gamma N_{I,j,v}, \\
\frac{dN_{I,j,v}}{dt} &= \alpha N_{E,j,v}, & \frac{dN_{R,j,v}}{dt} &= \gamma N_{I,j,v}, \\
\frac{dN_{i,E,v}}{dt} &= -\alpha N_{i,E,v}, & \frac{dN_{i,I,v}}{dt} &= -\gamma N_{i,I,v}, \\
\frac{dN_{i,I,v}}{dt} &= \alpha N_{i,E,v}, & \frac{dN_{i,R,v}}{dt} &= \gamma N_{i,I,v},
\end{aligned} \tag{3}$$

where: $i, j \in \{S, E, I, R\}$ and $v \in \{u, a, p, n\}$; $\alpha$ is the rate of progression from exposed to infectious states; and $\gamma$ is the recovery rate.

For a schematic of the model framework, see Supplementary Fig. 1.

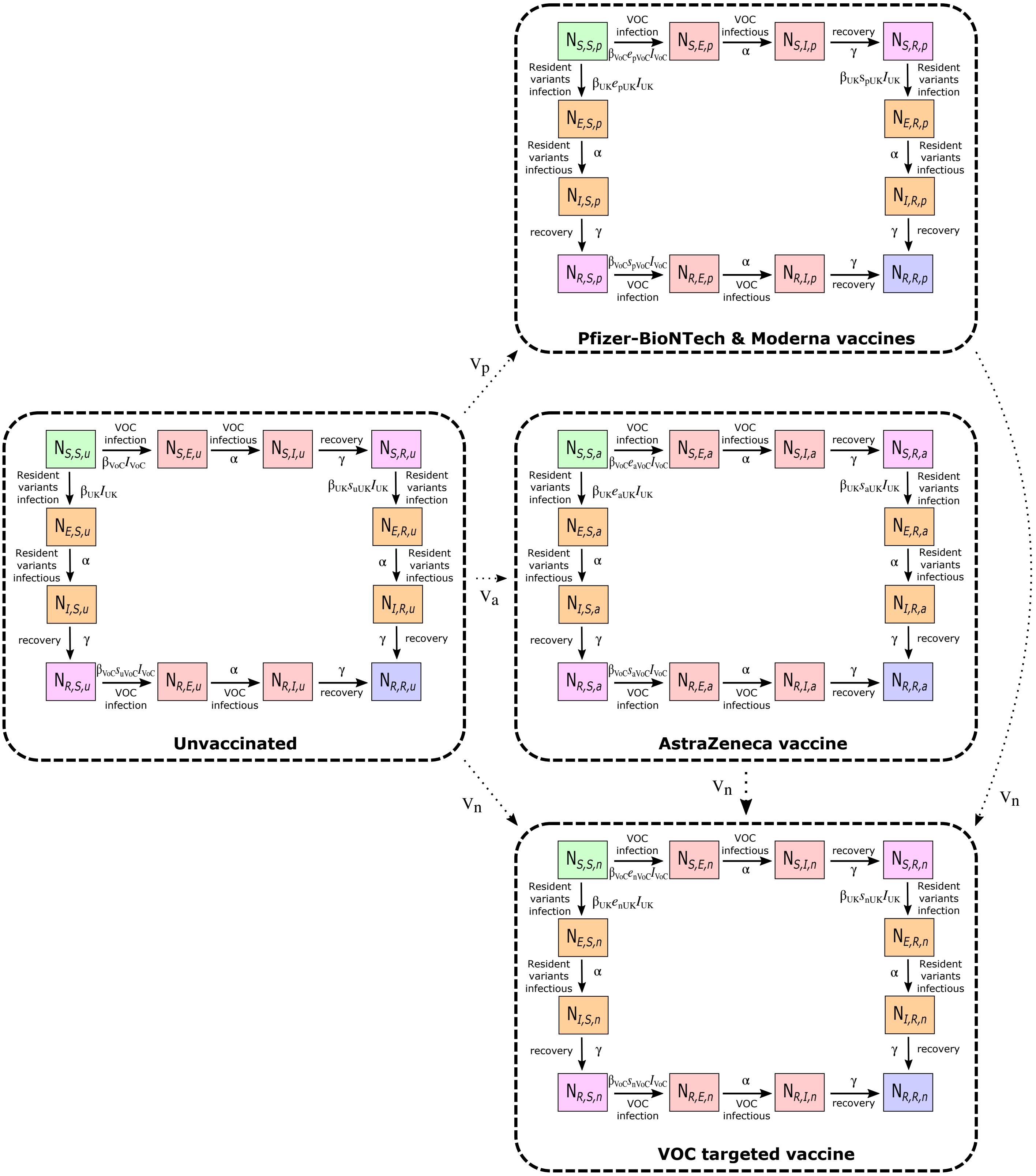

**Supplementary Figure 1: Schematic of our parsimonious SARS-CoV-2 transmission model consisting of an SEIR system for two variants and vaccination status.** We follow the populations given by $N_{i,j,v}$, where $i$ and $j$ denote whether the population is susceptible ($S$), exposed ($E$), infectious ($I$) or recovered ($R$) with respect to the resident variants and VOC, respectively, and $v$ denotes vaccination status as unvaccinated ($u$), or vaccinated with the AstraZeneca vaccine ($a$), Pfizer-BioNTech or Moderna vaccines ($p$), or an updated/new vaccine ($n$). The regions enclosed by the dashed lines depict the stratification of the population into four groups based on vaccination status: unvaccinated, Pfizer-BioNTech or Moderna vaccinated, AZ vaccinated and vaccinated with an updated/new vaccine. Boxes represent disease states (green: susceptible to both variants; orange: currently infected by resident variants; red: currently infected by VOC; pink: recovered from one variant, susceptible to the other variant blue: recovered from both variants). Solid arrows correspond to transitions between disease states. Dotted arrows correspond to transitions resulting from vaccination. We assumed unvaccinated susceptibles and recovereds could be vaccinated, corresponding to movements from the $N_{S,S,u}$, $N_{R,S,u}$, $N_{S,R,u}$ and $N_{R,R,u}$ states to the $N_{S,S,v}$, $N_{R,S,v}$, $N_{S,R,v}$ and $N_{R,R,v}$ states, respectively, at rate $V_v$ with $v \in \{a, p, n\}$. Dependent on the scenario, those previously vaccinated could subsequently receive the new/updated vaccine, corresponding to movements at rate $V_n$ from the $N_{S,S,\hat{v}}$, $N_{R,S,\hat{v}}$, $N_{S,R,\hat{v}}$ and $N_{R,R,\hat{v}}$ states (with $\hat{v} \in \{a, p)$ to the $N_{S,S,n}$, $N_{R,S,n}$, $N_{S,R,n}$ and $N_{R,R,n}$ states, respectively.

### 3 Age-structured SARS-CoV-2 transmission model

Here we present the basic model formulation that underpins the age-structured predictions of COVID-19 dynamics in England. We used a compartmental age-structured model, developed to simulate the spread of SARS-CoV-2 within the seven regions in England: East of England, London, Midlands, North East & Yorkshire, North West, South East and South West [20], with parameters inferred to generate a fit to deaths, hospitalisations, hospital occupancy and serological testing [21]. The model population is stratified by age, with the force of infection determined by the use of an age-dependent (who acquires infection from whom) social contact matrix for the UK [22, 23]. Additionally, we allow susceptibility and the probabilities of becoming symptomatic, being hospitalised and the risk of dying to be age dependent; these are matched to outbreak data for England. Finally, we account for the role of household isolation, by separating primary and secondary infections within a household (more details may be found in [20]). This allows us to capture household isolation by preventing secondary infections from playing a further role in onward transmission. Model parameters were inferred on a regional basis using regional time series of recorded daily hospitalisation numbers, hospital bed occupancy, ICU occupancy and daily deaths [21].

#### 3.1 Model description

The underlying system of equations accounts for the transmission dynamics, including symptomatic and asymptomatic transmission, household saturation of transmission and household quarantining. We describe the base model equations with respect to a single variant. To simulate the presence of a VOC alongside the existing resident variants, we duplicated the base model equations.

The population is stratified into multiple compartments: individuals may be susceptible ($S$), exposed ($E$), infectious with symptoms ($I$), or infectious and either asymptomatic or with very mild symptoms ($A$). Asymptomatic infections are assumed to transmit infection at a reduced rate given by $\tau$. To some extent, the separation into symptomatic ($I$) and asymptomatic ($A$) within the model is somewhat artificial as there are a wide spectrum of symptom severities that can be experienced.

We let superscripts denote the first infection in a household ($F$), a subsequent infection from a symptomatic household member ($SI$) and a subsequent infection from an asymptomatic household member ($SA$). A fraction ($H$) of the first detected cases (necessarily symptomatic) in a household are quarantined ($QF$), as are all their subsequent household infections ($QS$) - we ignore the impact of household quarantining on the susceptible population as the number in quarantine is assumed small compared with the rest of the population. The recovered class is not explicitly modelled, although it may become important once we have a better understanding of the duration of immunity. We omitted natural demography and disease-induced mortality in the formulation of the epidemiological dynamics.

We modelled two vaccination classes for individuals where it has been 14 days since they received their first and second dose of the vaccine; the 14-day delay allows partial immunity to develop (Supplementary Fig. 2). We included these within the $S$ and $E$ class by adding an additional vaccination subscript for the number of doses received; hence $S_{a,0}$ corresponds to susceptible unvaccinated individuals while $S_{a,2}$ corresponds to those that received their second dose of vaccine at least 14 days ago.

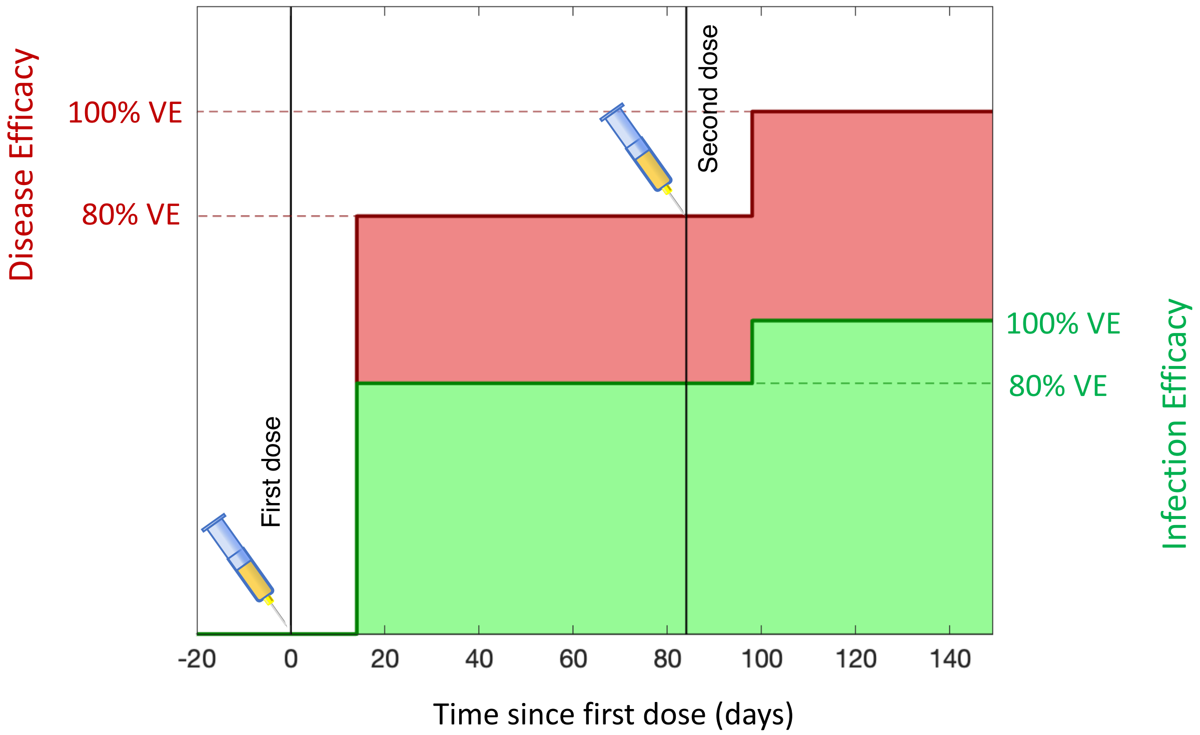

**Supplementary Figure 2: Dynamics of vaccine efficacy within an individual.** 14 days after the first dose partial efficacy is developed, and 14 days after the second dose efficacy is raised to its maximum value. We highlight two forms of efficacy: efficacy against severe symptomatic disease (red) which is captured by the parameter $z$ within the model; and protection against infection (green) which prevents all infection and acts on parameter $\sigma$.

The full equations are given by

$$
\begin{aligned}
\frac{dS_{a,0}}{dt} &= -\left(\lambda^F_{a,0} + \lambda^{SI}_{a,0} + \lambda^{SA}_{a,0} + \lambda^{Q}_{a,0}\right)\frac{S_{a,0}}{N_a} - V_{a,1}(t)S_{a,0}, \\
\frac{dS_{a,1}}{dt} &= V_{a,1}(t)S_{a,0} - \left(\lambda^F_{a,1} + \lambda^{SI}_{a,1} + \lambda^{SA}_{a,1} + \lambda^{Q}_{a,1}\right)\frac{S_{a,1}}{N_a} - V_{a,2}(t)S_{a,1}, \\
\frac{dS_{a,2}}{dt} &= V_{a,2}(t)S_{a,1} - \left(\lambda^F_{a,2} + \lambda^{SI}_{a,2} + \lambda^{SA}_{a,2} + \lambda^{Q}_{a,2}\right)\frac{S_{a,2}}{N_a} \\
\frac{dE^F_{1,a,v}}{dt} &= \lambda^F_{a,v}\frac{S_{a,v}}{N_a} - M\varepsilon E^F_{1,a,v}, \qquad v \in \{0,1,2\} \\
\frac{dE^{SI}_{1,a,v}}{dt} &= \lambda^{SI}_{a,v}\frac{S_{a,v}}{N_a} - M\varepsilon E^{SI}_{1,a,v}, \\
\frac{dE^{SA}_{1,a,v}}{dt} &= \lambda^{SA}_{a,v}\frac{S_{a,v}}{N_a} - M\varepsilon E^{SA}_{1,a,v}, \\
\frac{dE^{Q}_{1,a,v}}{dt} &= \lambda^{Q}_{a,v}\frac{S_{a,v}}{N_a} - M\varepsilon E^{Q}_{1,a,v}, \\
\frac{dE^{X}_{m,a,v}}{dt} &= M\varepsilon E^{X}_{m-1,a,v} - M\varepsilon E_{m,a,v}, \qquad X \in \{F, SI, SA, Q\} \\
\frac{dI^F_{a,v}}{dt} &= d_a(1-H)M\varepsilon E^F_{M,a,v} - \gamma I^F_{a,v}, \\
\frac{dI^{SD}_{a,v}}{dt} &= d_a M\varepsilon E^{SI}_{M,a,v} - \gamma I^{SI}_{a,v}, \\
\frac{dI^{SU}_{a,v}}{dt} &= d_a(1-H)M\varepsilon E^{SA}_{M,a,v} - \gamma I^{SA}_{a,v}, \\
\frac{dI^{QF}_{a,v}}{dt} &= d_a H M\varepsilon E^F_{M,a,v} - \gamma I^{QF}_{a,v}, \\
\frac{dI^{QS}_{a,v}}{dt} &= d_a H M\varepsilon E^{SA}_{M,a,v} + d_a\varepsilon E^{Q}_{a,v} - \gamma I^{QS}_{a,v}, \\
\frac{dA^F_{a,v}}{dt} &= (1-d_a)M\varepsilon E^F_{M,a,v} - \gamma A^F_{a,v}, \\
\frac{dA^S_{a,v}}{dt} &= (1-d_a)M\varepsilon (E^{SI}_{M,a,v} + E^{SA}_{M,a,v}) - \gamma A^S_{a,v}, \\
\frac{dA^Q_{a,v}}{dt} &= (1-d_a)M\varepsilon E^Q_{M,a,v} - \gamma A^Q_{a,v},
\end{aligned}
\tag{4}
$$

Here we have included $M$ latent classes. Throughout we have taken $M = 3$.

The forces of infection which govern the non-linear transmission of infection obey:

$$
\begin{aligned}
\lambda^F_{a,v} &= \beta_{nV}(t)\sigma_{a,v}\sum_{b,v}\left(I^F_{b,v} + I^{SI}_{b,v} + I^{SA}_{b,v} + \tau(A^F_{b,v} + A^S_{b,v})\right)\beta^N_{ba}, \\
\lambda^{SI}_{a,v} &= \beta_{nV}(t)\sigma_{a,v}\sum_{b,v} I^F_{b,v}\beta^H_{ba}, \\
\lambda^{SA}_{a,v} &= \beta_{nV}(t)\sigma_{a,v}\tau\sum_{b,v} A^F_{b,v}\beta^H_{ba}, \\
\lambda^{Q}_{a,v} &= \beta_{nV}(t)\sigma_{a,v}\sum_{b,v} D^{QF}_{b,v}\beta^H_{ba},
\end{aligned}
\tag{5}
$$

where $\beta^H$ represents household transmission and $\beta^N = \beta^S + \beta^W + \beta^O$ represents all other transmission locations, comprising school-based transmission ($\beta^S$), work-place transmission ($\beta^W$) and transmission in all other locations ($\beta^O$). These matrices are taken from Prem *et al.* [23] to allow easy translation to other geographic settings, although other sources such as POLYMOD [22] could be used. $\beta_{nV}(t)$ is a time-varying term that captures the increase in the B.1.1.7 variant in the UK and its generally higher rate of transmission (approximately 50% higher than the original). The temporal component of this term is derived from a higher-dimensional framework that models both the original and new variant as separate epidemic processes.

Two key parameters, together with the transmission matrix, govern the age-structured dynamics: $\sigma_a$ corresponds to the age-dependent susceptibility of individuals to infection (as we assume the probability associated with transmission from an infected individual to a susceptible contact to be constant with respect to age, we also absorb this quantity into the $\sigma_a$ parameters); $d_a$ the age-dependent probability of displaying symptoms (and hence potentially progressing to more severe disease). Both of these are also modified by the vaccine status, such that those that have received one or two doses of vaccine have a lower risk of infection and a lower risk of developing symptoms. The action of vaccine on the parameter $\sigma$ captures the protection against infection aspect of the vaccine, while the action on $d$ captures the efficacy against disease (Supplementary Fig. 2). We also define $\tau$ as the reduced transmission from asymptomatic infections compared to symptomatic infections; given the probability of displaying symptoms is less in the younger age groups, this parameter shapes the role of younger ages in onward transmission.

We require our model to capture both individual level quarantining of infected individuals and isolation of households containing identified cases. In a standard ODE framework this level of household structure is only achievable at large computational expense [24, 25]. Thus, we instead made a relatively parsimonious approximation to achieve a comparable effect.

We assume that all within household transmission originates from the first infected individual within the household (denoted with a superscript $F$, or $QF$ if they become quarantined). This allows us to assume that secondary infections within a household in isolation (denoted with a superscript $QS$ or $Q$) play no further role in any of the transmission dynamics. As a consequence, high levels of household isolation can drive the epidemic extinct, even if within household transmission is high – an effect not achievable with the standard SEIR-type modelling approach. This improved methodology also helps to capture to some degree household depletion of susceptibles (or saturation of infection), as secondary infections in the household are incapable of generating additional household infections.

For a schematic representation of the base model states and transitions (for a single variant), see Supplementary Fig. 3.

#### 3.2 Capturing social distancing

We obtained age-structured contact matrices for the United Kingdom from Prem et al. [23]. We used these contact matrices to provide information on normal levels of household transmission ($\beta^H_{ab}$, with the subscript $ab$ corresponding to transmission from age group $a$ against age group $b$), school-based transmission ($\beta^S_{ab}$), work-place transmission ($\beta^W_{ab}$) and transmission in all other locations ($\beta^O_{ab}$).

We assumed that any instigated non-pharmaceutical interventions (patterns of social-distancing or lockdown measures) leads to a reduction in the work, school and other matrices while increasing the strength of household contacts. Any given level of non-pharmaceutical interventions (NPIs), captured by the parameter $\phi$ between zero and one, therefore scales the four transmission matrices between their normal values (when $\phi = 0$) and their value under the most severe lockdown ($\phi = 1$).

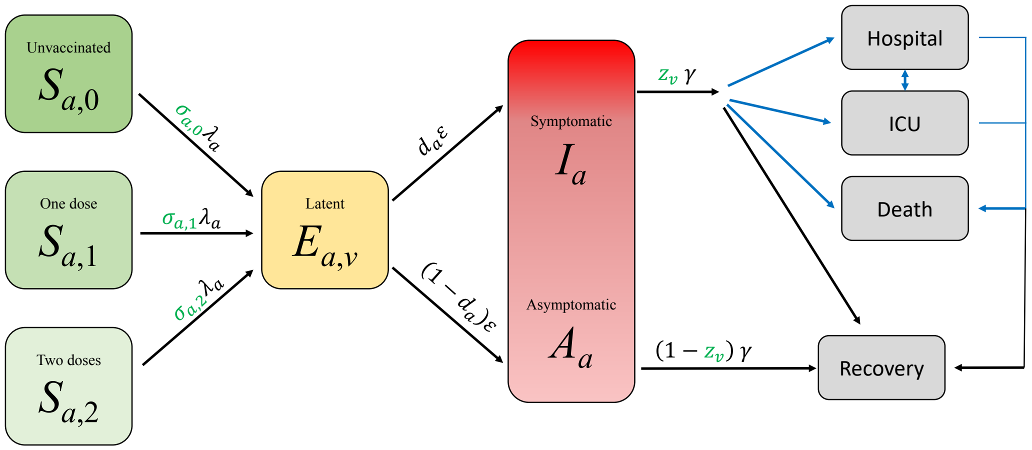

**Supplementary Figure 3: Representation of the basic model states and transitions for a single SARS-CoV-2 variant in the age-structured SARS-CoV-2 transmission model.** Black arrows show key epidemiological transitions while blue arrows show movements to observable states. Parameters in green show the action of vaccine on infection and probability of disease. In this graphic representation we have not separated the symptomatic and asymptomatic infections, to better illustrate that these compartments are somewhat ambiguous and depend on precise definitions being used at the time. We do not show stratification by first/secondary infection or quarantine states.

#### 3.3 Parameter Inference

As with any model of this complexity, there are multiple parameters that determine the dynamics. Some of these are global parameters, applying to all geographical regions, with others used to capture the regional dynamics. Some of these parameters are matched to the early outbreak data (including the resultant age-distribution of infection), however the majority are inferred by an MCMC process (Supplementary Table 2).

We would highlight that the parameters of $\alpha$ and $\tau$ are key in determining age-structured behaviour and are therefore essential in quantifying the role of school children in transmission [26]. We argue that a low $\tau$ and a low $\alpha$ are the only combination that are consistent with the growing body of data suggesting that levels of seroprevalence show only moderate variation across age-ranges [27], yet children are unlikely to display major symptoms, suggesting their role in transmission may be lower than for other respiratory infections [28, 29].

Throughout the pandemic, there has been noticeable heterogeneity between both the different regions of England and the devolved nations. In particular, London is observed to have a large proportion of early cases and a relatively steeper decline in the subsequent lockdown than the other regions and the devolved nations. We capture this heterogeneity in our model through the use of three regional parameters ($D_S^R$, $H_S^R$ and $I_S^R$), which act on the heterogeneous population pyramid of each region to generate key public health measurable observables (including hospital admissions and occupancy, ICU occupancy and deaths within 28-days of a positive COVID-19 test).

We inferred the regional strength of preventative behaviours, $\phi_R$, as a slowly varying parameter in the

MCMC scheme on a weekly basis.

**Supplementary Table 2:** Key model parameters and their source.

| Parameter | Description | Source |
|---|---|---|
| $V_{a,1}(t), V_{a,2}(t)$ | Time varying rate at which individuals in age group $a$ receive there first or second dose of vaccine | Assumptions based on UK vaccination supply |
| $\beta$ | Age-dependent transmission, split into household, school, work and other | Matrices from Prem *et al.* [23] |
| $\gamma$ | Recovery rate, changes with $\tau$, the relative level of transmission from undetected asymptomatics compared to detected symptomatics | Fitted from early age-stratified UK case data to match growth rate and $R_0$ |
| $d_{a,v}$ | Age-dependent and vaccine status dependent probability of displaying symptoms (and hence being detected), changes with $\alpha$ and $\tau$ | Fitted from early age-stratified UK case data to capture the age profile of infection. |
| $\sigma_{a,v}$ | Age-dependent and vaccine status dependent susceptibility, changes with $\alpha$ and $\tau$ | Fitted from early age-stratified UK case data to capture the age profile of infection. |
| $H^R$ | Household quarantine proportion $= 0.8\phi_R$ | Can be varied according to scenario |
| $N_a^R$ | Population size of a given age within each region | ONS |
| $\varepsilon$ | Rate of progression to infectious disease ($1/\varepsilon$ is the duration in the exposed class). $\varepsilon \sim 0.2$ | MCMC |
| $\alpha$ | Scales the degree to which age-structured heterogeneity is due to age-dependent probability of symptoms ($\alpha = 0$) or age-dependent susceptibility ($\alpha = 1$) | MCMC |
| $\tau$ | Relative level of transmission from asymptomatic compared to symptomatic infection | MCMC |
| $\phi^R$ | Regional relative strength of the lockdown restrictions; scales the transmission matrices. Can also be varied according to scenario. | MCMC |
| $\sigma^R$ | Regional modifier of susceptibility to account for differences in level of social mixing | MCMC |
| $E_0^R$ | Initial regional level of infection, rescaled from early age-distribution of cases | MCMC |
| $D_S^R$ | Regional scaling for the mortality probability $P_a$(Death\|Hospitalised)) | MCMC |
| $H_S^R$ | Regional scaling for the hospitalisation probability $P_a$(Hospitalised\|Symptomatic)) | MCMC |
| $I_S^R$ | Regional scaling for the ICU probability $P_a$(ICU\|Symptomatics)) | MCMC |

### 4 Stochastic invasion of a VOC and onward transmission

In the main text we described the use of a Gillespie simulation to model the initial phase of stochastic invasion, available at https://github.com/LouiseDyson/COVID19-variants-of-concern-modelling-paper. To cross-check the correctness of the simulation, we compared it with analytic results for a continuous-time multi-type branching process model with immigration, applying the general theory from [30] to our particular context.

#### 4.1 Model description

We considered the disease dynamics of an invading VOC into a population that has had previous exposure to a resident variant and has undergone an incomplete vaccination program with 3 different vaccines (Pfizer (Pf), AstraZeneca (AZ) and a new, VOC-targeted vaccine (N)). We assumed the novel variant to partially evade both infection-acquired and vaccine-acquired immunity.

We considered an epidemic in which the population was divided into 16 types, namely, 8 exposed types $E_{d,v}$ and 8 infectious types $I_{d,v}$ where $d \in \{\text{sus}, \text{rec}\}$ (for previous disease history) denotes whether an individual was susceptible to the current resident strain of COVID or had been infected previously and had recovered, and where $v \in \{\text{U}, \text{AZ}, \text{Pf}, \text{N}\}$ (for vaccination status) denotes whether an individual was unvaccinated or had been vaccinated with the AstraZeneca, Pfizer or VOC-targeted vaccine, respectively. We order the types in our model so that types 1-8 correspond to exposed cases and types 9-16 correspond to infectious cases; in both groups, the first four types are susceptible to the resident variant and the last four types recovered, and each of these four types have vaccination status ordered as listed above. Although there are 16 types in total, there are only 8 types-at-birth [31, Ch. 6], i.e. only 8 different types of individuals that have just been infected, corresponding to all possible combination of previous disease history with the resident variant and vaccination status. We denote by $f_j$ $(1 \leq j \leq 8)$ the fraction of the susceptible population of type-at-birth $j$, with $\sum_{j=1}^{8} f_j = 1$. Irrespective of their type-at-birth, exposed cases became infectious at a constant rate $\sigma$ and infectious cases had a constant recovery rate $\gamma$.

Susceptible individuals of type-at-birth $j$ had a relative susceptibility to the VOC, $\psi_j$, determined by their previous exposure to other variants and by their vaccination status. However, upon infection with the VOC, we assumed the transmissibility $\beta$ to be identical for each type-at-birth. Therefore, any individual infected with the VOC exerts a force of infection on all susceptible individuals of type-at-birth $j$ $(1 \leq j \leq 8)$ given by $\beta_j = c_j\beta$, where the constant $c_j$ scales $\beta$ by the relative proportions of type-at-birth $j$ in the population multiplied by the relative susceptibility of each type-at-birth to the VOC, i.e. $c_j = f_j\psi_j$.

Although the invasion phase of the epidemic is arguably more naturally modelled with a continuous-time birth-death process, this can be equivalently described with a continuous-time branching process, as explained in [30], where at each infection event, the infectious individual dies and is replaced by an identical copy of himself or herself and an exposed case of type-at-birth $j$. Following [30], we denote the lifetime (in this branching-process sense of time between any events) of a case of type $i$ by $\omega_i = \sigma$ for $1 \leq i \leq 8$ and by $\omega_i = \sum_{j=0}^{8} \beta_j + \gamma$ for $9 \leq i \leq 16$. The probability-generating function (pgf) for the offspring distribution of this process, i.e. the distribution of the number of cases of each type produced by a case of type $i$ at a single event, is given by:

$$P_i(\mathbf{s}) = \begin{cases} s_{i+8} & \text{for } 1 \leq i \leq 8 \\ \\ \frac{\sum_{j=0}^{8} \beta_j s_j s_i}{\omega_i} + \frac{\gamma}{\omega_i} & \text{for } 9 \leq i \leq 16 \end{cases} \tag{6}$$

where $\mathbf{s} = (s_i)_{i=1}^{16}$ is a (row) vector of length 16 whose components correspond to each different type of infected individual.

In our model, we initially considered a process that starts with a single case of type $i$ at time $t = 0$ and no other cases, and considered the vector $\mathbf{Y}_i(t) = (Y_{ij}(t))$ $(1 \leq i, j \leq 16)$, where the component $Y_{ij}(t)$ corresponds to the number of cases of type $j$ at time $t$. For each $i$, the random variable $\mathbf{Y}_i(t)$ has pgf

$$Q_i(t, \mathbf{s}) = \sum_{n_1, n_2, \ldots, n_{16}=0}^{\infty} \Pr(\mathbf{Y}_i(t) = \mathbf{n})\mathbf{s}^{\mathbf{n}}, \tag{7}$$

where both $\mathbf{s}$ and $\mathbf{n}$ are vectors, $\mathbf{s}^{\mathbf{n}}$ denotes the product $s_1^{n_1} \cdot \cdots \cdot s_{16}^{n_{16}}$ and $n_j$ is the number of cases of type $j$.

We obtain expressions for the $Q_i(t)$ by solving:

$$\frac{\partial Q_i(t, \mathbf{s})}{\partial t} = -\omega_i[Q_i(t, \mathbf{s}) - P_i(\mathbf{Q})], \quad \text{subject to } \mathbf{Q}(0, \mathbf{s}) = \mathbf{s}, \tag{8}$$

where $\mathbf{Q}(t, \mathbf{s}) = [Q_i(t, \mathbf{s})]_{i=1}^{16}$ is a vector. Setting $\mathbf{s} = \mathbf{0}$ gives the vector $\mathbf{q} = \mathbf{Q}(t, 0)$, the probability that, by time $t$, there are no cases of any type for a process starting with a single particle of type $i$.

We then considered a process that began with no cases of any type, but that allowed immigration of exposed cases of type $i$ at a constant rate $\eta_i$ and write $\eta = \sum_i \eta_i$. We denoted the total number of cases of all types at time $t$ for this process by $\mathbf{Z}(t) = (Z_j(t))$ $(1 \leq j \leq 16)$, with the associated generating function

$$R(t, \mathbf{s}) = \sum_{n_1, n_2, \ldots, n_{16}=0}^{\infty} \Pr(\mathbf{Z}(t) = \mathbf{n})\mathbf{s}^{\mathbf{n}}, \tag{9}$$

where $\mathbf{n} = (n_1, n_2, \ldots, n_{16})$ as before. We obtain $R(t, \mathbf{s})$ by solving:

$$\frac{\partial R(t, \mathbf{s})}{\partial t} = -\eta R(t, \mathbf{s}) + \sum_{i=0}^{16} \eta_i R(t, \mathbf{s}) Q_i(t, \mathbf{s}), \quad \text{subject to } R(0, \mathbf{s}) = 1. \tag{10}$$

Setting $\mathbf{s} = \mathbf{0}$ as before, we find the probability of zero cases of any type at time $t$, $r(t) = R(t, 0)$. This should no longer be interpreted as a probability that the outbreak of the VOC has gone extinct, as immigration allows new cases to enter the population even when the total number of cases is zero. However, we can loosely interpret $\lim_{t\to\infty} 1 - r(t)$ as the probability that a VOC is successfully established in the population and that the resulting epidemic grows exponentially.

#### 4.2 Calculation of means

The vector of the mean number of cases of each type at time $t$, $\mathbf{m}(t) = \mathbb{E}[\mathbf{Z}(t)] = (\mathbb{E}[Z_j(t)])_{j=1}^{16}$ is given by:

$$\mathbf{m}(t) = \mathbf{m}(0)\mathrm{e}^{t\Omega} + \int_0^t \eta \mathrm{e}^{(t-\tau)\Omega} \mathrm{d}\tau \tag{11}$$

given by $\Omega_{i,j} = \frac{\partial P_i}{\partial s_j}(\mathbf{1}) - \omega_i$.

##### 4.3 Calculation of variances

In order to obtain the variance matrix for the random vector $\mathbf{Z}(t) = (Z_j(t))$, representing the number of cases of each type $j$ at time $t$, we follow the general theory in [30] applied to our particular case and consider the variance matrix $W_i(t)$ for $(1 \leq i \leq 16)$ for an outbreak that began with no cases and accounts only for immigration of cases of type $i$ at a constant rate $\eta_i$, which can be calculated via:

$$W_i(t) = \int_0^t \eta_i \mathbb{E}[\mathbf{Y}_T(t)^* \mathbf{Y}_T(t) | T = \tau] \, \mathrm{d}\tau, \tag{12}$$

for $1 \leq i \leq 16$, where $\mathbf{Y}_T(t)$ represents the vector of the number of cases of each type generated by a single initial case of type $i$ that appeared in the population via immigration at a random time $T$. If the matrix $\Omega$ is diagonalisable, so that $\Omega = ADA^{-1}$, we have that:

$$W_i(t) = \eta_i H \Delta H^{-1} \text{Vec}(C) \tag{13}$$

where the matrix $H = A \otimes (A^*)^{-1} \otimes (A^*)^{-1}$, Vec is the operator that vectorises a matrix by stacking its columns and $C$ is the $16^2 \times 16$ matrix consisting of block $16 \times 16$ matrices $C_i$ given by:

$$C_i = \omega_i \Big( \frac{\partial^2 P_i}{\partial s_j^2}(\mathbf{1}) + \text{diag}(\frac{\partial P_i}{\partial s_j}(\mathbf{1})) + e_i^* e_i - e_i^* \frac{\partial P_i}{\partial s_j}(\mathbf{1}) - \frac{\partial P_i}{\partial s_j}(\mathbf{1})^* e_i \Big) \tag{14}$$

for $1 \leq i \leq 16$. Here, $e_i$ refers to the standard $i^{th}$ basis (row) vector. In (13), $\Delta$ is the diagonal matrix whose $(i, j, k)^{th}$ entry is given by:

$$\frac{1}{\delta_i - (\delta_j + \delta_k)} \left[ \frac{\mathrm{e}^{\delta_i t} - 1}{\delta_i} - \frac{\mathrm{e}^{(\delta_j + \delta_k)t} - 1}{\delta_j + \delta_k} \right], \tag{15}$$

where $\delta_i$ is the $i^{th}$ diagonal entry of the matrix $D$.

Finally, the variance-covariance matrix of the random vector $\mathbf{Z}(t) = (Z_j(t))$ is given by

$$\text{Var}[\mathbf{Z}(t)] = \sum_{i=1}^{16} W_i(t), \tag{16}$$

and therefore the variance of the random number of total infected cases at time $t$ is obtained as

$$\text{Var}\left[ \sum_{j=1}^{16} Z_j(t) \right] = \sum_{r=1}^{16} \sum_{s=1}^{16} \text{Var}[\mathbf{Z}(t)]_{(r,s)}, \tag{17}$$

i.e. the sum all of the entries of the variance-covariance matrix of $\mathbf{Z}(t)$.

##### 4.4 Consideration of deterministic approximation

We finally considered the number of cases required for the ratio of the mean number of cases to the standard deviation to be constant over time. We interpret this quantity as the width of the confidence region around the mean number of cases over time, which after a certain time should grow proportionally to the mean curve. For different scenarios, we plot the mean number of cases over time along with the standard deviation divided by the mean (see Supplementary Fig. 4), and note that the latter quantity reaches a steady state in all scenarios by the time 100 cases have been infected, on average. In our main analysis, we seed our deterministic models with 2,000 infected cases on the 17th May 2021, which clearly satisfies the necessary conditions for a deterministic approximation to be valid.

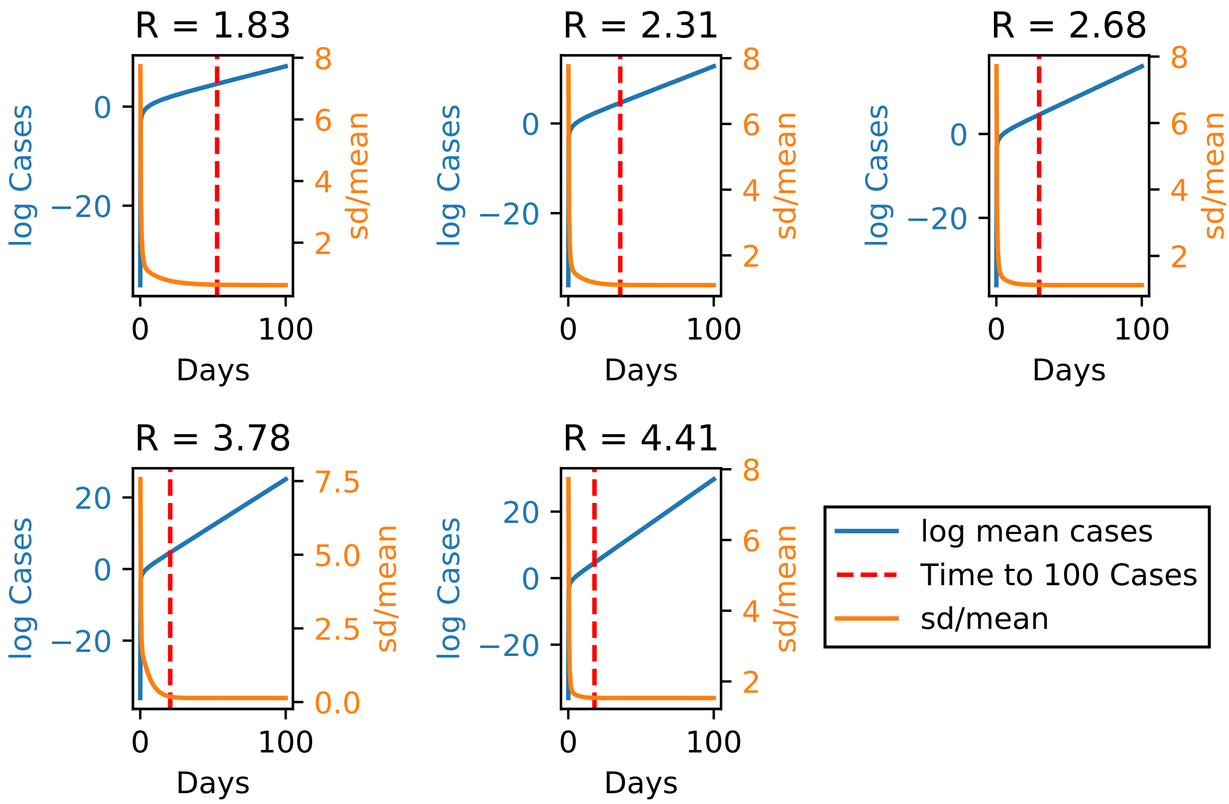

**Supplementary Figure 4:** Plot of the log of the expected number of cases over time (blue curve) for different values of R for the invading VOC, as well as the standard deviation of the number of cases divided by the mean (orange curve). By the time the expected total number of cases has reached 100 (dashed red line), the standard deviation divided by the mean is approximately constant over time. We conclude that, once this has happened, the number of cases is approximately normally distributed over time, centred around the mean, and hence a deterministic approximation of the epidemic is appropriate.

### 5 Additional figures

#### 5.1 Parsimonious SARS-CoV-2 transmission model

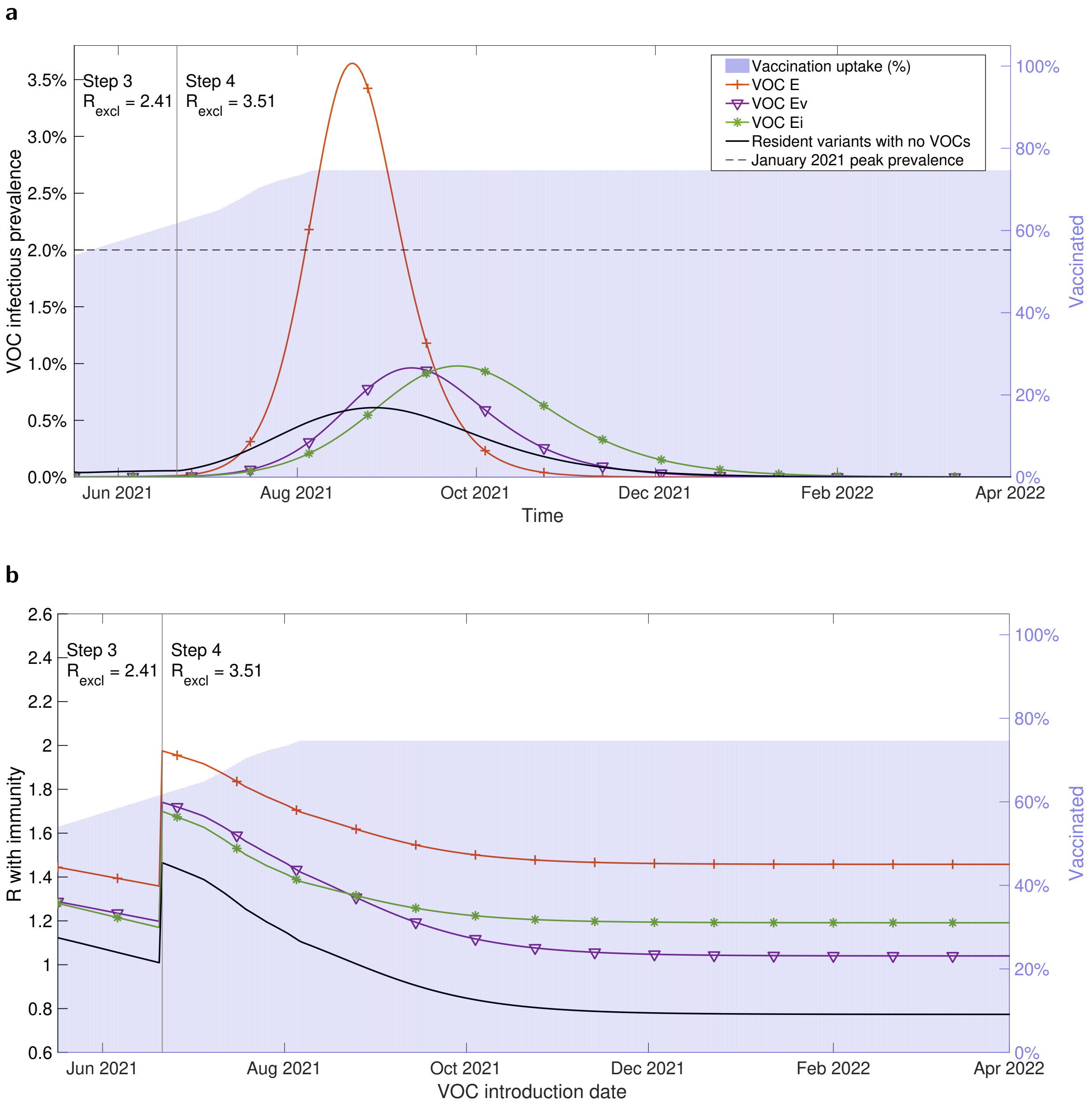

**Supplementary Figure 5: Sensitivity of epidemiological measures to VOCs Ev and Ei.** In all simulations we seeded 2,000 VOC infecteds on 17th May 2021. Both VOC Ev and VOC Ei had the same transmissibility as the resident variants. VOC Ev (purple solid line with inverted triangle markers) had immune escape to vaccination, whereas VOC Ei (green solid line with asterisk markers) had immune escape to natural infection. For reference, we display the timeseries for VOC E (same transmissibility, with immune escape to both vaccination and natural infection, depicted by the orange line with plus sign markers) and for the resident variants in the absence of any VOC being introduced (black solid line). We represent the vaccine uptake in the population through time via background shading, the transition time into Step 4 of the relaxation roadmap by the vertical solid line and we state the assumed R excluding immunity values for the resident variants ($R_{\text{excl}}$) throughout Steps 3 and 4, respectively. **a** Infectious prevalence timeseries for the VOC. **b** R with immunity (y-axis) with respect to the date of a VOC being introduced (x-axis). For the 'Resident variants with no VOCs' scenario the displayed profile corresponds to the instantaneous R with immunity of the resident variants.

**a**

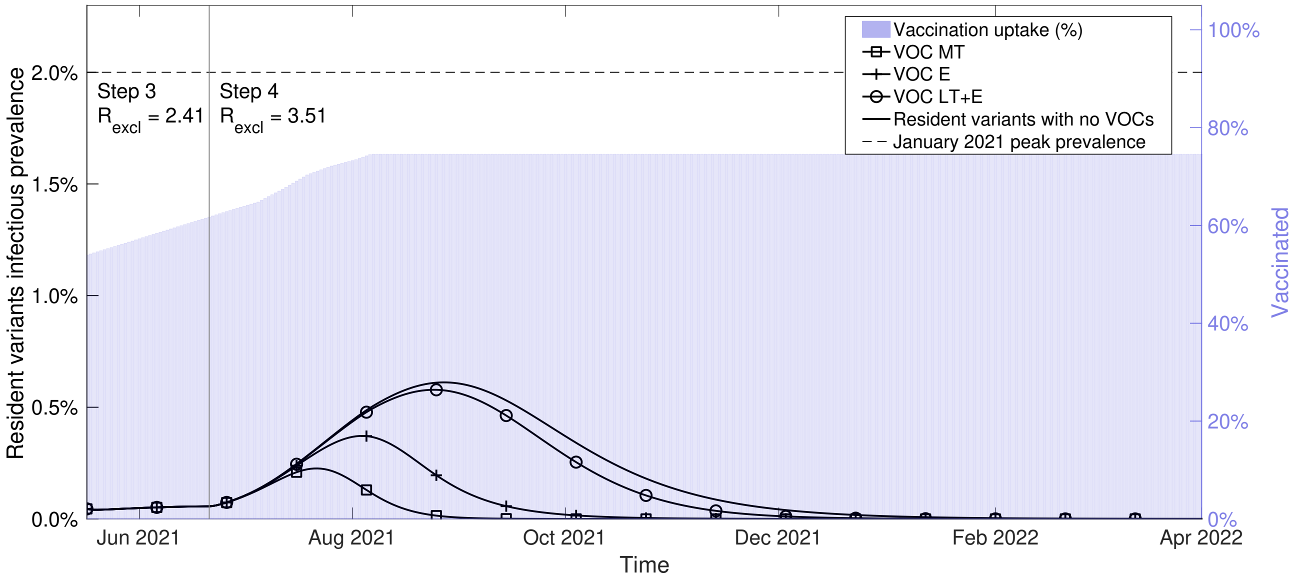

**b**

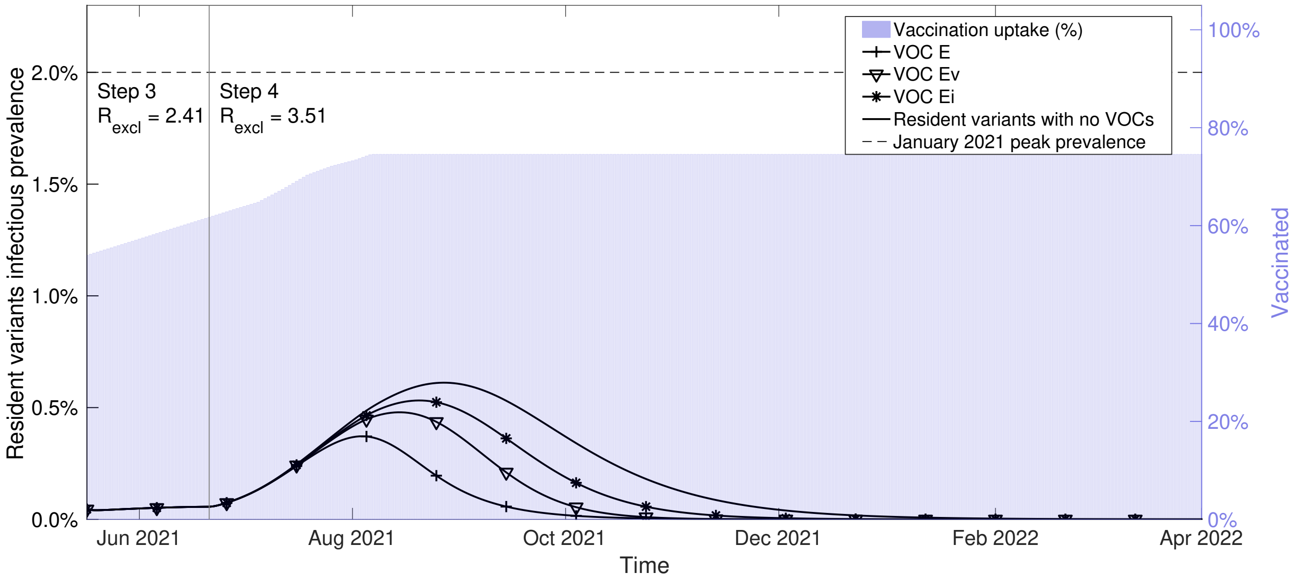

**Supplementary Figure 6: Resident variants infectious prevalence over time under each putative VOC scenario.** We present infectious prevalence timeseries for the resident variants when the 2,000 infecteds of the specified VOC were introduced on 17th May 2021: **a** VOC MT (square markers), VOC E (plus sign markers), VOC LT+E (circle markers); **b** VOC Ev (inverted triangle markers) had immune escape to vaccination, VOC Ei (asterisk markers) had immune escape to natural infection. We also display in panel **b** the timeseries for VOC E (same transmissibility, with immune escape to both vaccination and natural infection, depicted by the line with plus sign markers). In both panels we show the infectious prevalence for the resident variants in the absence of any VOC being introduced (line with no markers), represent the vaccine uptake in the population through time via background shading, depict the transition time into Step 4 of the relaxation roadmap by the vertical solid line and state the assumed R excluding immunity values for the resident variants ($R_{\text{excl}}$) throughout Steps 3 and 4, respectively.

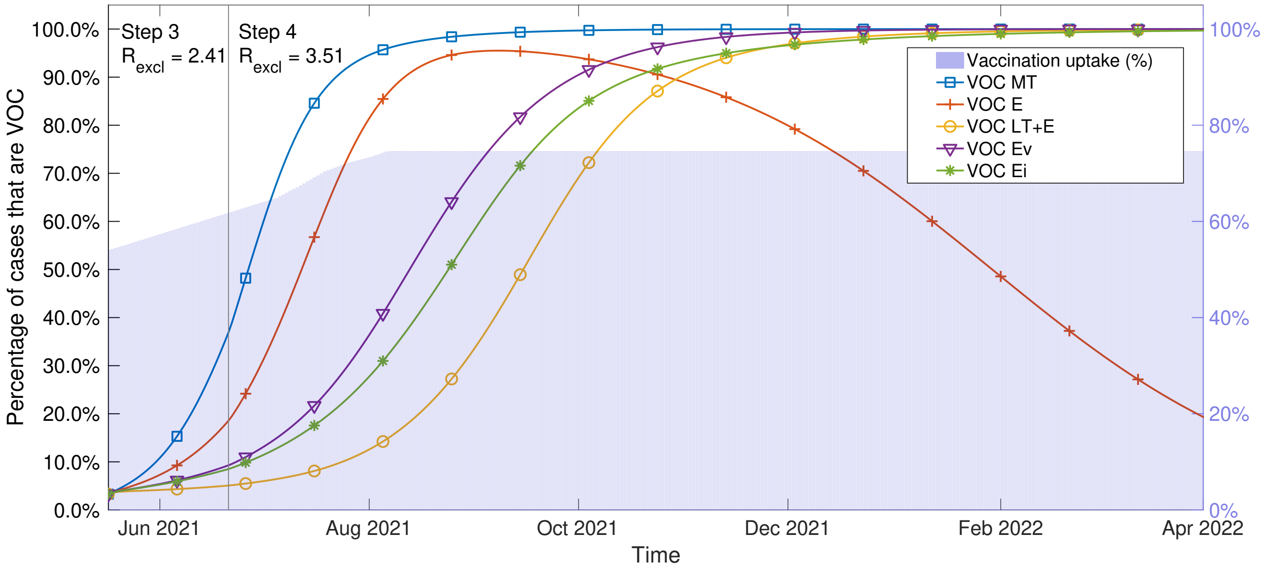

**Supplementary Figure 7: Percentage of cases attributed to the VOC (versus resident variants) over time under each putative VOC scenario.** We present timeseries for the percentage of cases attributable to the VOC when 2,000 infecteds of the specified VOC were introduced on 17th May 2021: VOC MT (square markers); VOC E (plus sign markers); VOC LT+E (circle markers); VOC Ev (inverted triangle markers); or VOC Ei (asterisk markers). We represent the vaccine uptake in the population through time via background shading, depict the transition time into Step 4 of the relaxation roadmap by the vertical solid line and state the assumed R excluding immunity values for the resident variants ($R_{\text{excl}}$) throughout Steps 3 and 4, respectively. We highlight that the apparent reversal in the VOC E scenario to resident variants becoming predominant again results from the declines in the waves of infection for the VOC and resident variants coinciding with different relative rates of decrease. We also note that infectious prevalence in scenario VOC E is very low for both the VOC and resident variants moving into 2022.

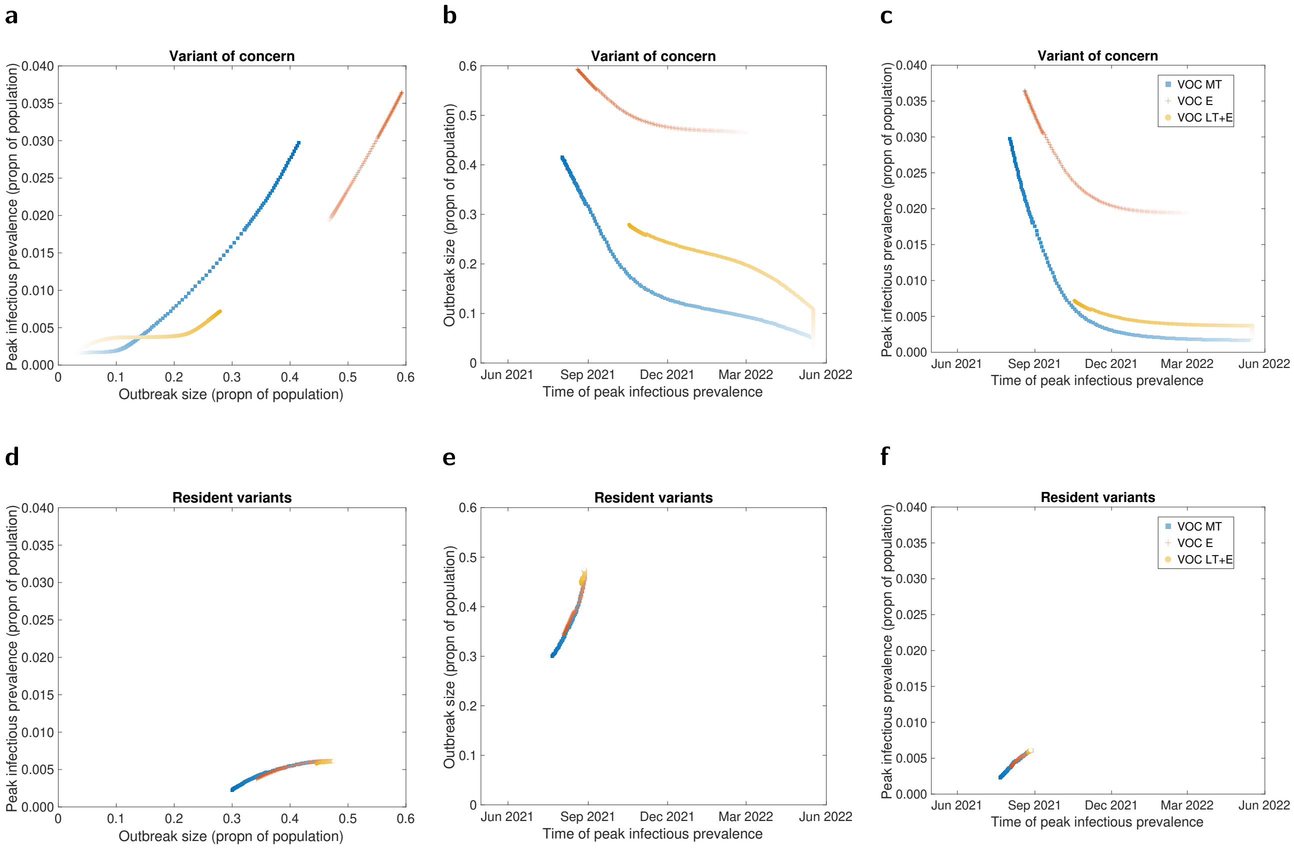

**Supplementary Figure 8: Relationship between outbreak size, peak in infectious cases and timing of the peak in infectious cases.** For each VOC scenario, we generated a series of 92 data points, one for each tested day of introducing the given VOC into the population. Our VOC introduction dates spanned 17th May 2021 to 1st November 2021 inclusive (depicted in the data points by a transition from dark marker shading to light marker shading). We display outputs for VOCs (panels **a-c**), and resident variants (panels **d-f**).

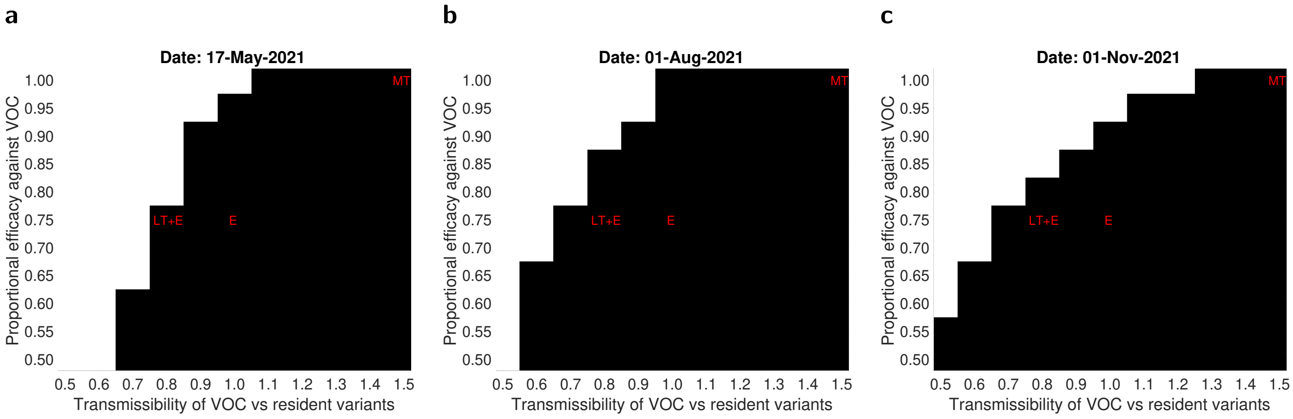

**Supplementary Figure 9: Sensitivity of the relative effective R (R with immunity) to the date of introduction of seed VOC infecteds and epidemiological characteristics of the VOC.** We present simulations performed using the following introduction dates for 2,000 VOC infecteds: **(a)** 17th May 2021; **(b)** 1st August 2021; **(c)** 1st November 2021. We identify the combinations of relative transmissibility of the VOC ($T_r$) and proportional efficacy against the VOC for which $R_{eff}^{VOC} \geq 1$ and the relative effective R for the VOC versus the resident variants $(R_{eff}^{VOC}/R_{eff}^{\text{res}}) \geq 1$. Shaded regions correspond to the parameter sets satisfying the stated criteria. Glyph labels denote the parameter combinations used for our three main illustrative VOCs: VOC MT, VOC E and VOC LT+E.

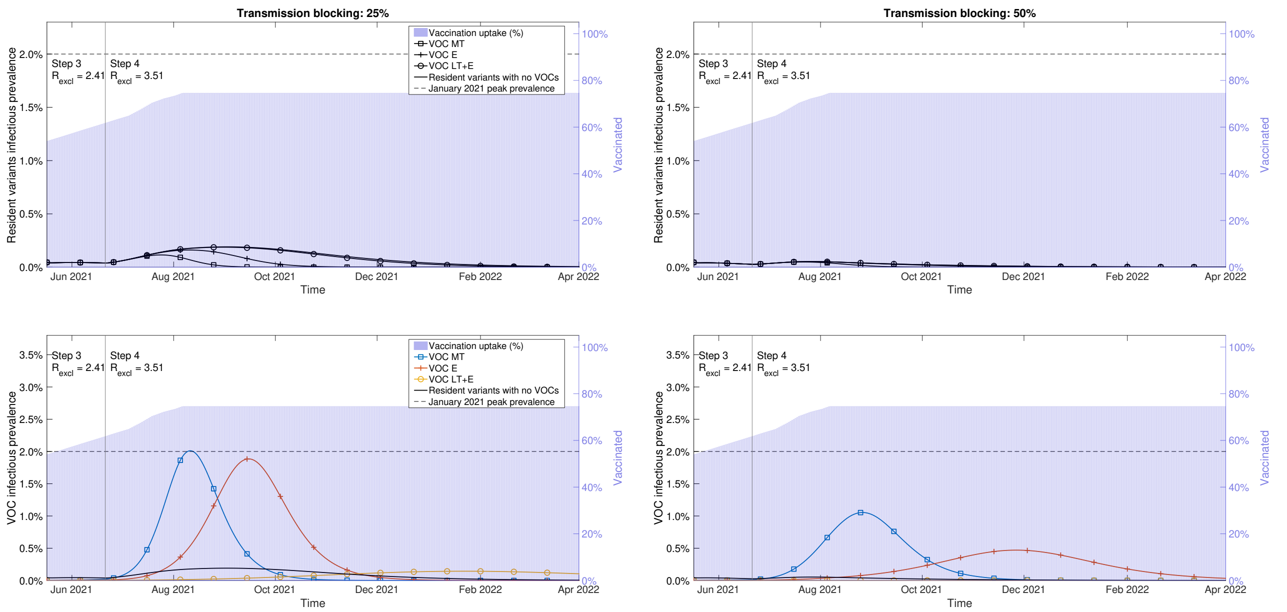

**Supplementary Figure 10: Temporal infectious timeseries for scenarios including a reduction in onward transmission (transmission blocking) action of immunity.** In all simulations we seeded 2,000 VOC infecteds on 17th May 2021. We simulated scenarios with immunity (provided from either prior infection or vaccination) resulting in a reduction in transmissibility of 25% (left column) and 50% (right column). We display temporal profiles for resident variants infectious (top row) and VOC infectious (bottom row). We display the infectious prevalence over time for our illustrative VOC scenarios: more transmissible (VOC MT, lines with square markers); equal transmissibility with vaccine immune escape (VOC E, lines with plus sign markers); less transmissible with vaccine immune escape (VOC LT+E, lines with circle markers). We also display the infectious prevalence for the resident variants in the absence of any VOC being introduced (black solid line with no markers) and the background shading corresponds to the vaccine uptake in the population through time.

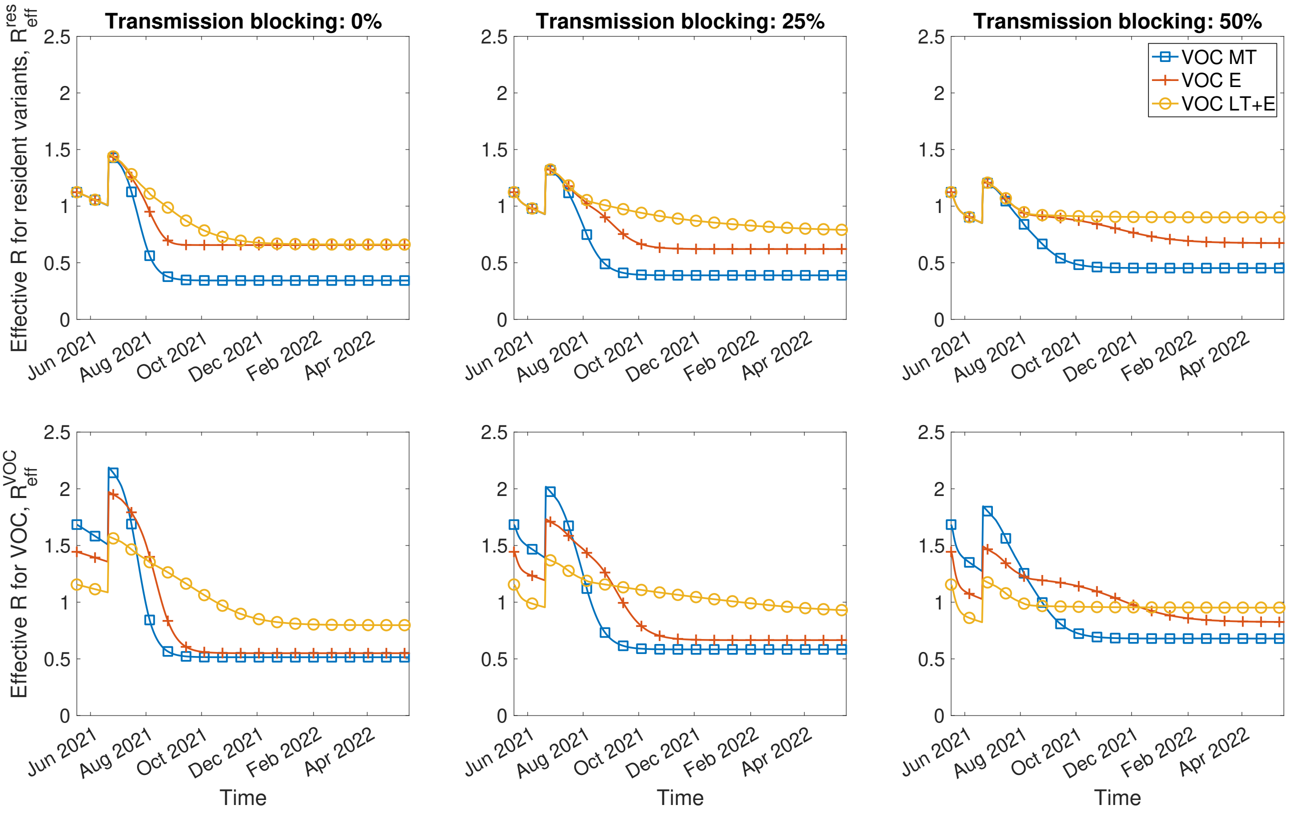

**Supplementary Figure 11: Sensitivity of effective R (R with immunity) to different assumptions for the reduction in onward transmission (transmission blocking) action of immunity.** In each panel we present results when introducing into the infectious disease dynamics either VOC MT (blue solid line, square markers), VOC E (orange dashed line, plus sign markers) or VOC LT+E (yellow dotted line, circle markers). We seed 2,000 initial VOC infecteds on 17th May 2021. The top row gives the effective R for the resident variants, $R_{\text{eff}}^{w}$. The bottom row gives the effective R for the VOC, $R_{\text{eff}}^{\text{VOC}}$. We simulated scenarios with immunity (provided from either prior infection or vaccination) resulting in a reduction in transmissibility of 0% (left column), 25% (middle column) and 50% (right column).

#### 5.2 Age-structured SARS-CoV-2 transmission model

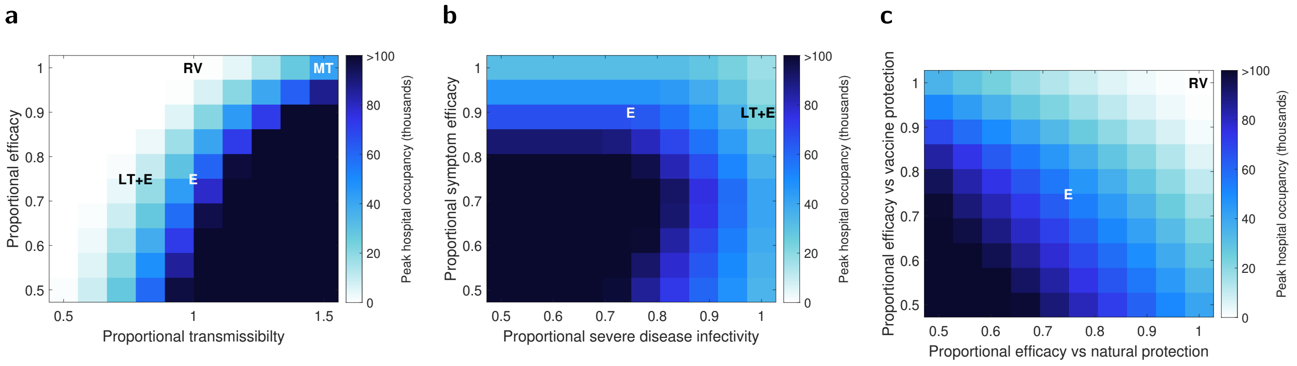

**Supplementary Figure 12: Variation in peak hospital occupancy due to a VOC for a range of different VOC characteristics.** Each panel shows mean values for peak hospital occupancy across 50 simulations in which the roadmap is run to completion. Specific VOC characteristics considered elsewhere are indicated for reference by labelled letters. **a** Proportional transmissibilty to resident variants versus proportional efficacies (proportional to the efficacy towards the resident variants) against infection and hospitalisation (severe disease) for both naturally acquired protection and vaccination. We fixed efficacy against symptoms at 90%. **b** Proportional efficacies against developing symptoms versus efficacy against hospitalisation (severe disease) for both naturally acquired protection and vaccination. We fixed infection efficacy at 75% and set the transmissibility of the VOC equal to resident variants. **c** Proportional efficacies against infection and hospitalisation (severe disease) for naturally acquired protection versus vaccination. We fixed the efficacy against symptoms at 90% and set the transmissibility of the VOC equal to resident variants.

#### 5.3 VOC-targeted vaccines

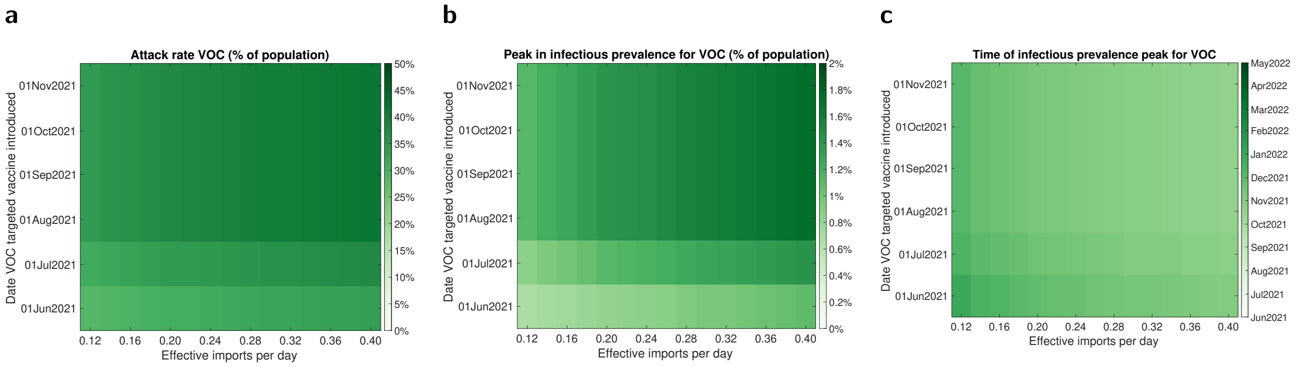

**Supplementary Figure 13: Sensitivity of epidemic trajectories to the introduction time of a VOC targeted vaccine for VOC E, using a prioritisation scheme with unvaccinated individuals given precedence followed by those who had received one of the pre-existing vaccines.** We performed simulations using the parsimonious SARS-CoV-2 transmission model for differing effective VOC importation counts and introduction date of a VOC targeted vaccine and evaluated the following epidemiological summary statistics for the resultant VOC outbreak: **(a)** final size; **(b)** peak in infectious prevalence; **(c)** time of peak in infectious prevalence.
